# Efficacy and Safety of CAR T-cell Therapy in Relapsed/Refractory B-cell Acute Lymphoblastic Leukemia: A Systematic Review and Meta-analysis

**DOI:** 10.1101/2025.08.31.25334789

**Authors:** Rakhshanda khan, Sai Manohar Raju Obbani, Sanjana Thota, Sri Mani Krishna Satvik Matta, Anisha Rafique Dodhia, Kalyan Naik Gugulothu, Yashas Maragowdanahalli Somegowda, Harsimran kaur, Mridula Joginapally, Akshaya Junnuthula, Harshawardhan Dhanraj Ramteke, Syeda Hafsa Noor-Ain, Dr. Manish Juneja

## Abstract

**Introduction:** Acute lymphoblastic leukemia (ALL) is the most common childhood cancer, with rising global incidence. The prognosis for patients with relapsed or refractory B-cell ALL (r/r B-ALL) remains poor, necessitating novel therapies. Chimeric Antigen Receptor T-cell (CAR T-cell) therapy has shown promise in treating r/r B-ALL, offering significant improvements in remission rates.

**Methods:** A comprehensive literature search was conducted across PubMed, Embase, and Cochrane Library for studies evaluating CAR T-cell therapy in r/r B-ALL. Randomized controlled trials, cohort, and case-control studies were included, focusing on efficacy and safety outcomes. Data were extracted and pooled using random-effects models. The risk of bias was assessed with the New Ottawa Scale.

**Results:** A total of 32 studies were included, involving 1,55,365 patients. The pooled relapse rate was 0.39 (95% CI: 0.29–0.49), with no significant difference between CD19 and CD22 CAR T-cell therapies (p = 0.88). Co-stimulatory agents like 4-1BB showed the most favorable relapse rate of 0.38 (95% CI: 0.27–0.49). The overall Cytokine Release Syndrome (CRS) rate was 0.63 (95% CI: 0.53–0.73), and neurotoxicity occurred at a rate of 0.32 (95% CI: 0.24–0.41).

**Conclusion:** CAR T-cell therapy is effective in treating r/r B-ALL, with high remission rates and manageable adverse events. The choice of co-stimulatory agent and antigen target influences relapse outcomes. Further research is needed to refine CAR T-cell constructs and optimize patient-specific treatments.

## Introduction

Acute lymphoblastic leukemia (ALL) is a hematologic malignancy characterized by the rapid proliferation of immature lymphocytes, predominantly of B-cell lineage. It stands as the most prevalent form of pediatric cancer, accounting for approximately 25% of all childhood cancers [1]. The global incidence of ALL has been on the rise, with an increase of 59.06% from 1990 to 2021, culminating in 168,879 new cases in 2021 alone [2]. This upward trend is particularly pronounced in low socio-demographic index regions, where the incidence of leukemia cases has shown a clear upward trajectory across all age groups from 1990 to 2019 [3].

Despite advancements in treatment, the prognosis for patients with relapsed or refractory B-cell ALL (r/r B-ALL) remains dismal. In children, the 5-year overall survival rate plummets to approximately 20% upon relapse, while adults fare even worse, with survival rates below 40% due to high-risk features and limited treatment options [4]. Traditional therapies, including chemotherapy and hematopoietic stem cell transplantation, often fail to achieve durable remissions in these patient populations, underscoring the urgent need for novel therapeutic strategies.

Chimeric Antigen Receptor T-cell (CAR T-cell) therapy has emerged as a transformative approach in the treatment of r/r B-ALL. This immunotherapeutic modality involves the genetic modification of a patient’s T cells to express receptors specific to tumor-associated antigens, thereby enhancing the immune system’s ability to target and eliminate malignant cells. The FDA’s approval of tisagenlecleucel (Kymriah) in 2017 marked a significant milestone, offering a new horizon for pediatric and young adult patients with r/r B-ALL [5].

Clinical trials have demonstrated promising efficacy of CAR T-cell therapies, with complete remission rates reaching up to 90% in certain cohorts [6]. However, these therapies are not without their challenges. Adverse events such as cytokine release syndrome (CRS) and immune effector cell-associated neurotoxicity syndrome (ICANS) are notable concerns, necessitating careful patient monitoring and management [6]. Moreover, the high cost of CAR T-cell therapies, often exceeding $500,000 per infusion, poses significant barriers to accessibility and widespread adoption, particularly in low-resource settings [7].

The complexity and cost of CAR T-cell therapies highlight the need for ongoing research to optimize their efficacy and safety profiles. Investigating alternative antigen targets, such as CD22 and dual-target approaches like CD19/CD22, may offer avenues to overcome issues related to antigen escape and enhance therapeutic outcomes [8]. Additionally, exploring costimulatory domains, such as CD28 and 4-1BB, could further refine CAR T-cell constructs to improve both efficacy and safety [9].

This systematic review and meta-analysis aim to critically evaluate the current evidence on the efficacy and safety of CAR T-cell therapies in the treatment of r/r B-ALL. By synthesizing data from multiple studies, we seek to provide comprehensive insights into the therapeutic potential of CAR T-cell therapy, identify factors influencing treatment outcomes, and inform clinical decision-making to enhance patient care in this challenging disease context.

## Methods

### Literature Search

A comprehensive search was conducted across PubMed, Embase, Cochrane Library, and ClinicalTrials.gov using terms like “CAR T-cell therapy,” “relapsed/refractory B-ALL,” “efficacy,” and “safety.” Inclusion criteria focused on clinical trials and studies reporting efficacy and adverse events, with no language restrictions. This process followed the PRISMA (Preferred Reporting Items for Systematic Reviews and Meta-Analyses) guidelines [10], ensuring thorough and transparent search methods. The Protocol was registered with Prospero with number CRD420251101772.

### Study Selection and Data Extraction

Studies were selected based on predefined inclusion criteria, including randomized controlled trials, cohort studies, and case-control studies that evaluated CAR T-cell therapy in patients with relapsed or refractory B-cell acute lymphoblastic leukemia (r/r B-ALL). Studies must report on efficacy (e.g., complete remission, survival rates) and safety (e.g., cytokine release syndrome, neurotoxicity) outcomes. Two independent reviewers conducted the selection process using a systematic approach to ensure all relevant studies were considered. Any discrepancies were resolved through consensus or a third reviewer. Data were extracted using a standardized form, capturing study characteristics (e.g., author, year, sample size), patient demographics (e.g., age, sex), treatment protocols (e.g., CAR T-cell constructs, dosages), and primary outcomes (e.g., remission rates, adverse events). Statistical analysis was performed to compute pooled estimates of efficacy and safety outcomes using random-effects models.

### Risk of Bias Assessment

The risk of bias in included studies was assessed using the New Ottawa Scale, evaluating selection, comparability, and outcome domains [11]. Studies were scored based on the clarity of participant selection, control of confounders, and the reliability of outcome measures. Discrepancies were resolved through consensus or a third reviewer.

### Statistical Analysis

Statistical analyses were conducted using Stata 18.0. For ordinal rating scales, data were treated as continuous variables, with mean changes from baseline reported. The treatment effect was represented as the mean difference (MD) with 95% confidence intervals (CI). When standard deviations (SD) were not provided, these were calculated from p-values, in line with the guidelines from the Cochrane Handbook for Systematic Reviews of Interventions.

To assess heterogeneity, both the Chi² test and the I² statistic were employed. A significance threshold of P < 0.10 was used for the Chi² test, while an I² value greater than 50% indicated substantial heterogeneity. In cases of low heterogeneity (I² < 50%), a fixed-effects model was used to calculate pooled estimates of mean differences between the intervention groups. If significant heterogeneity was present, a random-effects model was applied.

Subgroup analysis was performed based on various factors and targeted brain regions. Sensitivity analysis using a leave-one-out approach was conducted to assess the influence of individual studies on the overall results in the presence of significant heterogeneity.

## Results

### Demographics

A total of 1356 studies were analyzed, out of which 678 were later removed because of duplicates, rest 200 were removed because of ineligibility, furthermore 198 were removed for other reasons. Total of 280 records were screened and 157 were excluded, only 32 were included [12–42]. The process of screening is depicted in Prisma flow diagram, figure 1. The total number of patients in the meta-analysis were 155365, out of which 78974 were males and 76252 were females. The treatment group had 155206 patients and control group had 78 patients. The average follow up was of 32.5 ± 7 months, whereas, Average age of the population was 17.11 years and dosage was 1.98 x 10^6. The treatment approaches included in this study are as follows: **Autologous CD19-CAR T-cells**, with 5 patients, **Blinatumomab bridging CAR-T**, involving 18 patients, and **Brexucabtagene autoleucel**, which was administered to 121 patients. **CAR T-Cells** were used in a total of 154,638 cases, while **CD19 CART** was applied to 64 patients. Additionally, **CD22-CAR T** therapy was provided to 55 patients, **KTE-C19** was used for 11 patients, and **KTE-X19** was administered to 24 patients. Lastly, **Obecabtagene autoleucel** was used in 127 cases, and **Tisagenlecleucel** was given to 124 patients.

**Figure 1.**
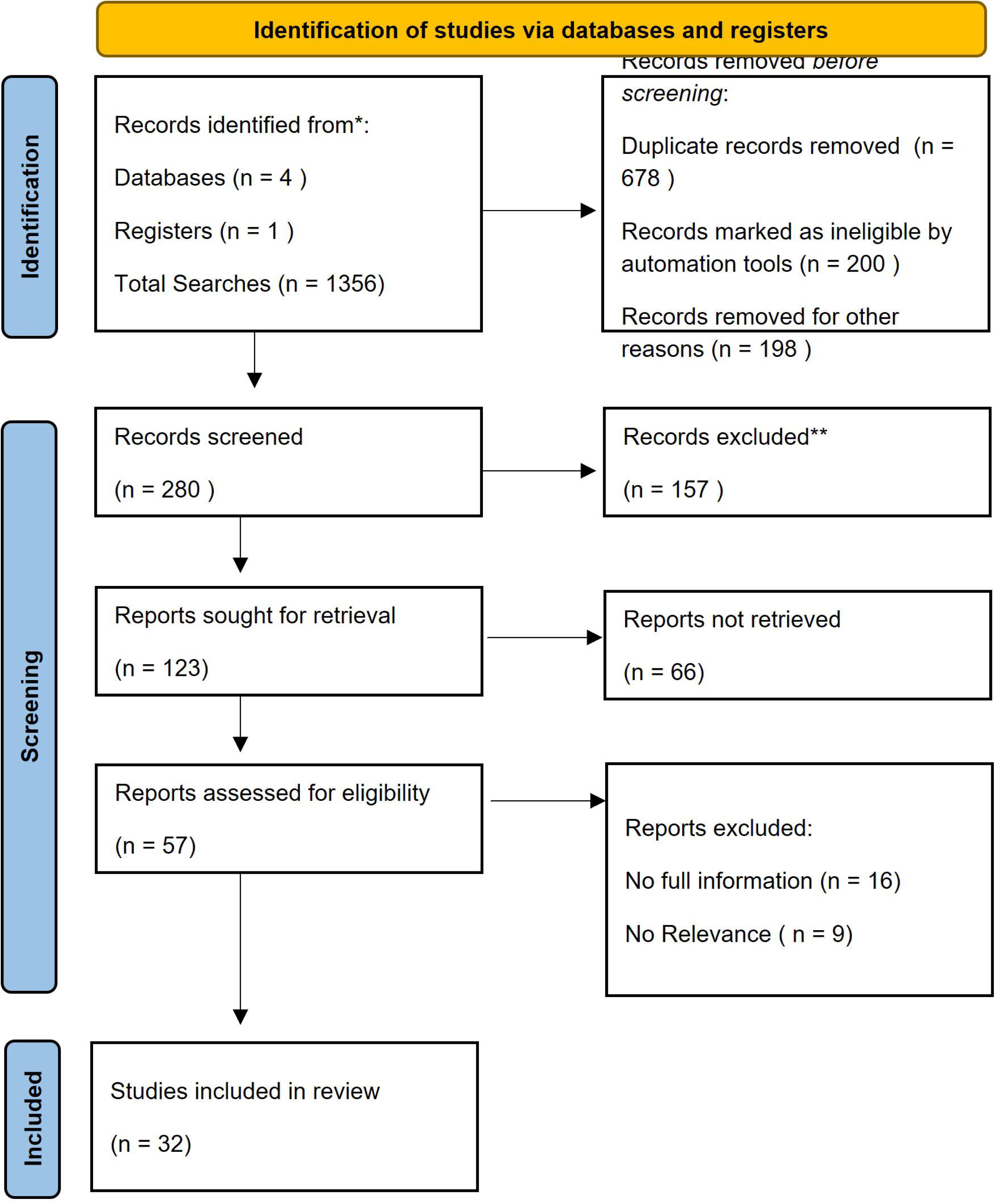
Prisma Flow Diagram

### Comparison of Relapse Rates in CD19 and CD22 CAR T-Cell Therapies: A Subgroup Analysis

The forest plot provides a comprehensive analysis of the relapse rates in patients receiving CD19 and CD22 CAR T-cell therapies, comparing multiple studies. For CD19, studies such as **Shah et al. (2025)** show a relatively high relapse rate of 0.63 (95% CI: 0.41–0.85) and contribute significantly to the overall analysis with a weight of 3.79%. In contrast, **Hirama et al. (2019)** report a much lower relapse rate of 0.25 (95% CI: 0.00– 0.55), with a weight of 2.74%, highlighting variability across different studies. Other studies like **Levine et al. (2021)** and **Ma et al. (2019)** also present varying relapse rates, with the former at 0.62 (95% CI: 0.41–0.85) and the latter at 0.40 (95% CI: 0.10–0.70), showing significant heterogeneity in the reported outcomes.

On the other hand, **CD22 CAR T-cell studies** exhibit a different trend. For instance, **Fry et al. (2018)** report a relapse rate of 0.52 (95% CI: 0.31–0.74), while **Minnema et al. (2024)** show a slightly higher rate of 0.48 (95% CI: 0.35–0.61), both contributing notably to the overall analysis. In contrast, **Pan et al. (2019)** report a much lower relapse rate of 0.12 (95% CI: 0.01–0.23) but with a relatively smaller weight in the overall analysis, indicating variability in the results across different studies within the CD22 category.

When combining both CD19 and CD22 studies, the overall pooled relapse rate is 0.39 (95% CI: 0.29–0.49), with moderate heterogeneity (I² = 9.62%), suggesting consistency in relapse rates across both treatment strategies. The test for group differences (Q(1) = 0.03, p = 0.88) shows no significant difference between the two therapies, indicating that both CD19 and CD22 CAR T-cell therapies have similar relapse outcomes, despite individual study variations. The overall conclusion points to comparable efficacy in preventing relapse with both treatment strategies, although variations in relapse rates among different studies underscore the influence of specific factors within each study’s design. Figure 2 A.

**Figure 2. A.**
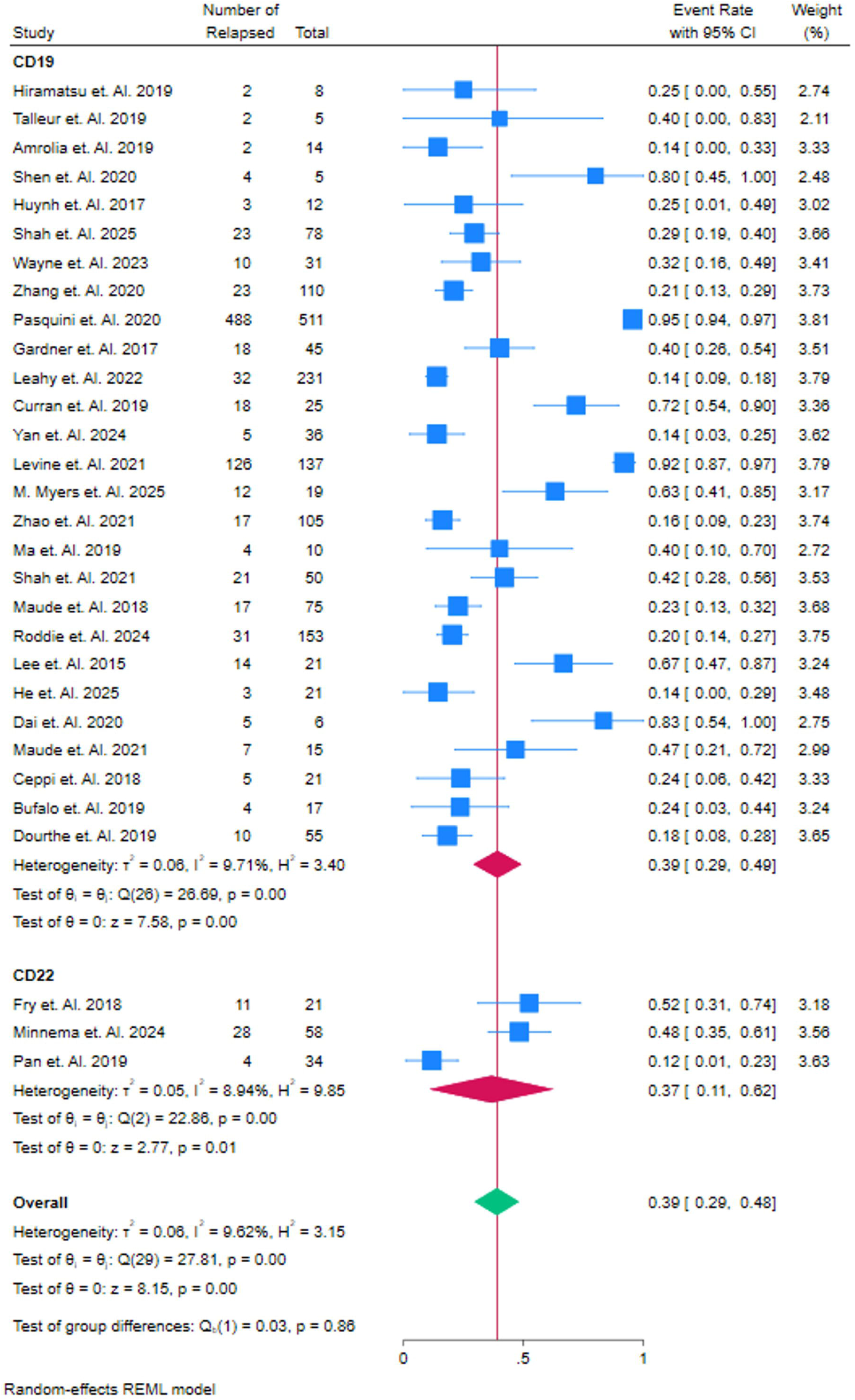
Event rate of patients who had Relapsed, Sub Group Analysis of Domain

### Comparison of Relapse Rates Across Co-Stimulatory Agents in CAR T-Cell Therapy

The forest plot presents a detailed analysis of the event rates of relapse in patients treated with various co-stimulatory agents, including 4-1BB, CD137, CD19, CD28, and CD3F. In the **4-1BB** group, studies such as **Pasquini et al. (2020)** show a relatively high relapse rate of 0.95 (95% CI: 0.91–0.97), with a substantial weight of 3.81%, whereas **Talleur et al. (2019)** report a much lower rate of 0.40 (95% CI: 0.00–0.83), with a weight of 2.11%. The overall pooled relapse rate for 4-1BB is 0.38 (95% CI: 0.27–0.49), reflecting moderate variability in the outcomes. For the **CD137** co-stimulatory agent, **Hiramatsu et al. (2019)** report a relapse rate of 0.25 (95% CI: 0.00–0.55) with a weight of 2.74%, and the overall pooled relapse rate for CD137 is consistent at 0.25 (95% CI: 0.05–0.55), suggesting limited variability across studies. In the **CD19** group, **Yan et al. (2024)** report a relapse rate of 0.14 (95% CI: 0.03–0.25) with a weight of 3.62%, indicating a relatively low event rate compared to other agents. The **CD28** co-stimulatory group demonstrates more variation, with **Wayne et al. (2023)** showing a relapse rate of 0.32 (95% CI: 0.16–0.49) and **Shah et al. (2021)** reporting 0.42 (95% CI: 0.28–0.56), leading to an overall pooled event rate of 0.45 (95% CI: 0.24–0.66). Finally, **CD3F**, as evaluated by **Shen et al. (2020)**, shows the highest relapse rate of 0.80 (95% CI: 0.45–1.00) with a weight of 2.48%, reflecting its higher relapse risk compared to the other agents. The overall analysis suggests that while CD19 and CD3F co-stimulatory agents show relatively favorable outcomes, there is notable variability in the efficacy of co-stimulatory agents in CAR T-cell therapies. The differences in relapse rates underscore the importance of selecting the appropriate co-stimulatory molecule to optimize therapeutic outcomes. Figure 2B.

**Figure 2 B.**
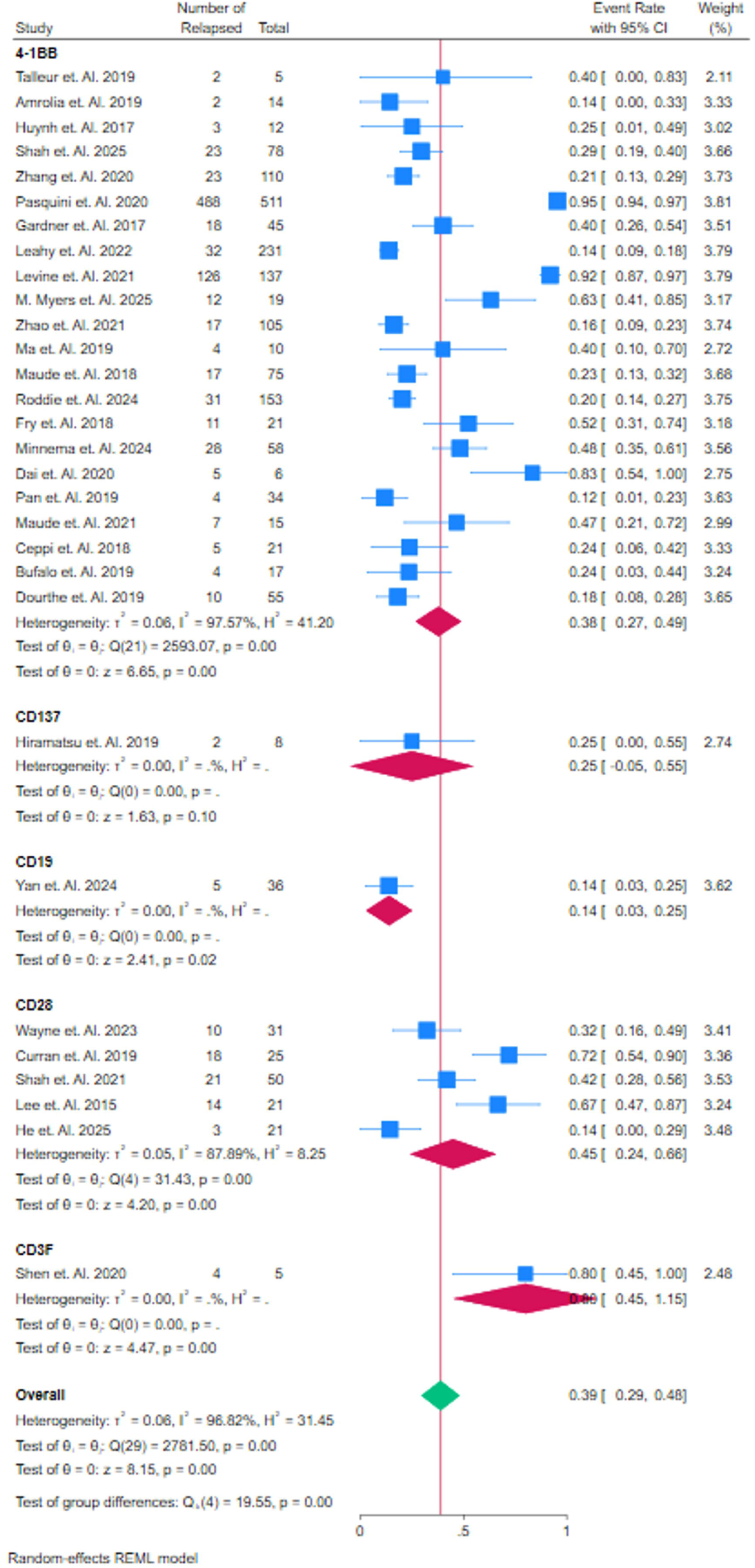
Event rate of patients who had relapsed, Sub group analysis of Co-Stimulatory Agent

### Comparison of Relapse Rates in CAR T-Cell Therapy Alone vs. CAR T-Cell Therapy Combined with HSCT

The forest plot presents a detailed comparison of relapse rates in patients treated with **CAR T-cell therapy (CART)** alone versus **CAR T-cell therapy combined with Hematopoietic Stem Cell Transplantation (HSCT)**. In the **CART alone** group, studies such as **Pasquini et al. (2020)** show a notably high relapse rate of 0.95 (95% CI: 0.91–0.97), contributing a substantial weight of 3.81% to the overall analysis, while **Dourthe et al. (2019)** reports a significantly lower relapse rate of 0.18 (95% CI: 0.08–0.28), with a weight of 3.65%. This results in an overall pooled relapse rate for **CART alone** of 0.33 (95% CI: 0.21–0.46), indicating notable variability across studies. On the other hand, the **CART + HSCT** group, which combines CAR T-cell therapy with stem cell transplantation, shows a mixed set of results. For example, **Shah et al. (2025)** reports a relapse rate of 0.29 (95% CI: 0.19–0.40), with a weight of 3.66%, while **Levine et al. (2021)** reports a higher relapse rate of 0.62 (95% CI: 0.41–0.85), contributing 3.79% to the pooled analysis. The overall relapse rate for **CART + HSCT** is 0.39 (95% CI: 0.29–0.48), showing a slightly higher relapse rate compared to **CART alone**.

Despite the individual variability within the studies, the pooled analysis reveals that there is no significant difference between the two therapies, as indicated by the non-significant test for group differences (Q(1) = 1.42, p = 0.23). The relapse rates for **CART alone** and **CART + HSCT** are similar, with the former at 0.33 (95% CI: 0.21–0.46) and the latter at 0.39 (95% CI: 0.29–0.48). Therefore, both treatment strategies appear to offer comparable efficacy in preventing relapse, suggesting that adding HSCT to CAR T-cell therapy does not provide a significant improvement in relapse rates. Figure 2C.

**Figure 2. C.**
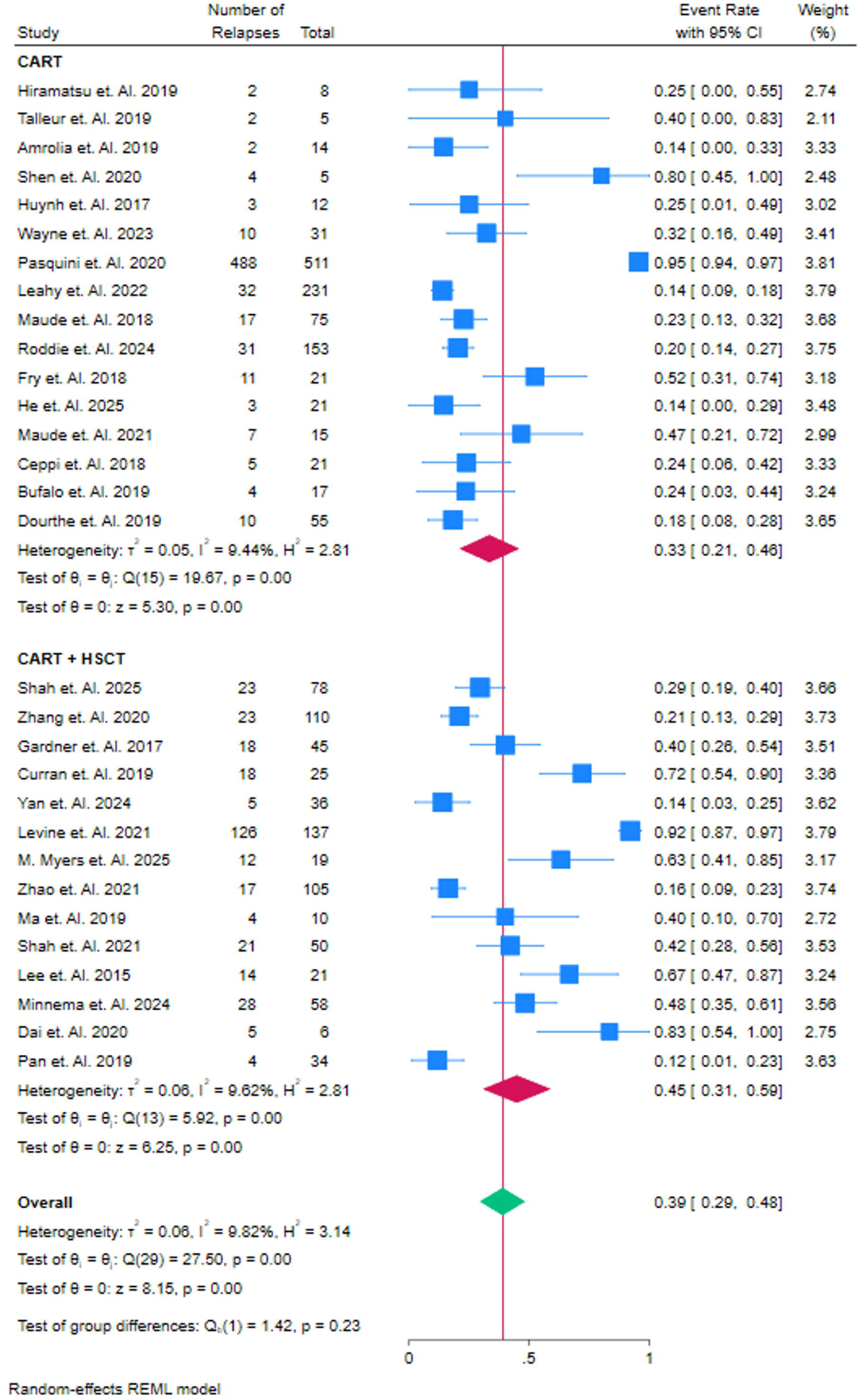
Event rate of patients who had relapsed, Sub group analysis of CART + HSCT therapy or HSCT alone therapy

### Comparison of Complete Remission Rates in CD19 and CD22 CAR T-Cell Therapies

The forest plot provides a comparison of complete remission rates in patients treated with **CD19** and **CD22** CAR T-cell therapies. In the **CD19** group, studies such as **Shah et al. (2025)** report a high remission rate of 0.74 (95% CI: 0.65–0.84), contributing significantly to the overall pooled remission rate, while **Hirama et al. (2019)** report a lower rate of 0.38 (95% CI: 0.04–0.71), reflecting a wider variability. The study by **Pasquini et al. (2020)** shows the highest remission rate of 0.95 (95% CI: 0.91–0.97), with a weight of 3.69%. The overall pooled remission rate for **CD19** is 0.69 (95% CI: 0.61–0.77), highlighting its effectiveness in inducing complete remission in patients.

In the **CD22** group, **Fry et al. (2018)** report a remission rate of 0.52 (95% CI: 0.31–0.74), contributing 3.01% to the overall analysis, while **Minnema et al. (2024)** show a slightly lower remission rate of 0.41 (95% CI: 0.20– 0.54), with a weight of 3.53%. The study by **Pan et al. (2019)** reports the highest remission rate in this group, 0.71 (95% CI: 0.55–0.88), with a weight of 3.38%. The pooled remission rate for **CD22** is 0.67 (95% CI: 0.60– 0.75), which is slightly higher than the **CD19** group.

While **CD22** therapies show a marginally higher overall remission rate (0.67) compared to **CD19** therapies (0.69), the test for group differences (Q(1) = 2.01, p = 0.16) indicates that this difference is not statistically significant. Both **CD19** and **CD22** CAR T-cell therapies are effective in achieving complete remission, with high rates across studies, suggesting their comparable efficacy in treating relapsed or refractory B-cell leukemia. Figure 3A.

**Figure 3. A.**
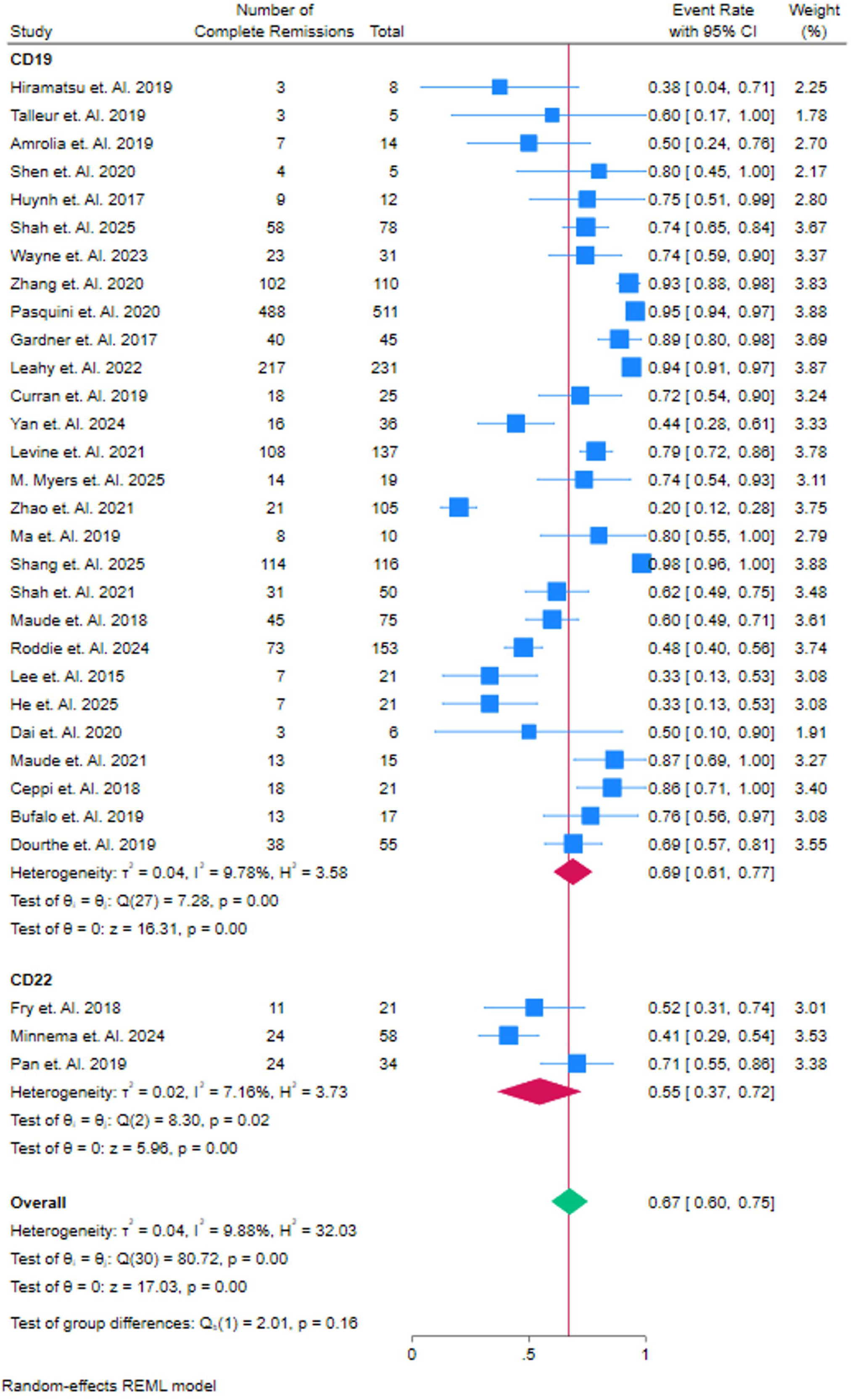
Event rate of patients who had Complete Remission, Sub Group Analysis of CD Target

### Comparison of Relapse Rates Across Co-Stimulatory Agents in CAR T-Cell Therapy

The forest plot provides a comparison of relapse rates in patients treated with different co-stimulatory agents, including **4-1BB**, **CD137**, **CD19**, **CD28**, and **CD3F**. In the **4-1BB** group, studies such as **Shah et al. (2025)** report a high event rate of relapse at 0.93 (95% CI: 0.88–0.98), contributing significantly to the overall pooled event rate. Other studies, such as **Talleur et al. (2019)**, show a relatively lower relapse rate of 0.60 (95% CI: 0.17–1.00), suggesting variability in the effectiveness of this co-stimulatory agent. The pooled relapse rate for **4-1BB** is 0.72 (95% CI: 0.63–0.80), indicating moderate efficacy across studies. For **CD137**, **Hiramatsu et al. (2019)** report a lower relapse rate of 0.38 (95% CI: 0.04–0.71), with a relatively low weight, leading to a pooled relapse rate of 0.38 (95% CI: 0.04–0.71), showing limited variability and lower efficacy in preventing relapse.

In the **CD19** group, **Yan et al. (2024)** report a relapse rate of 0.44 (95% CI: 0.28–0.61), which contributes 3.33% to the overall analysis. The pooled relapse rate for **CD19** is 0.44 (95% CI: 0.28–0.61), reflecting relatively consistent outcomes across studies. The **CD28** group, represented by **Wayne et al. (2023)**, reports a higher relapse rate of 0.74 (95% CI: 0.59–0.90), leading to a pooled relapse rate of 0.56 (95% CI: 0.39–0.73), suggesting a moderate risk of relapse compared to **CD19** and **CD137**. In the **CD3F** group, **Shen et al. (2020)** report a relatively higher relapse rate of 0.80 (95% CI: 0.45–1.00), with a pooled relapse rate of 0.67 (95% CI: 0.60–0.75), indicating a higher relapse rate compared to other co-stimulatory agents. Overall, the pooled relapse rate across all co-stimulatory agents is 0.67 (95% CI: 0.60–0.75), with moderate heterogeneity (I² = 46.88%). The test for group differences (Q(4) = 12.41, p = 0.01) indicates significant variability between the agents, with **4-1BB** showing the highest efficacy in reducing relapse rates, while **CD137** demonstrates lower efficacy. This suggests that different co-stimulatory agents have varying degrees of effectiveness in preventing relapse in CAR T-cell therapy. Figure 3B.

**Figure 3. B.**
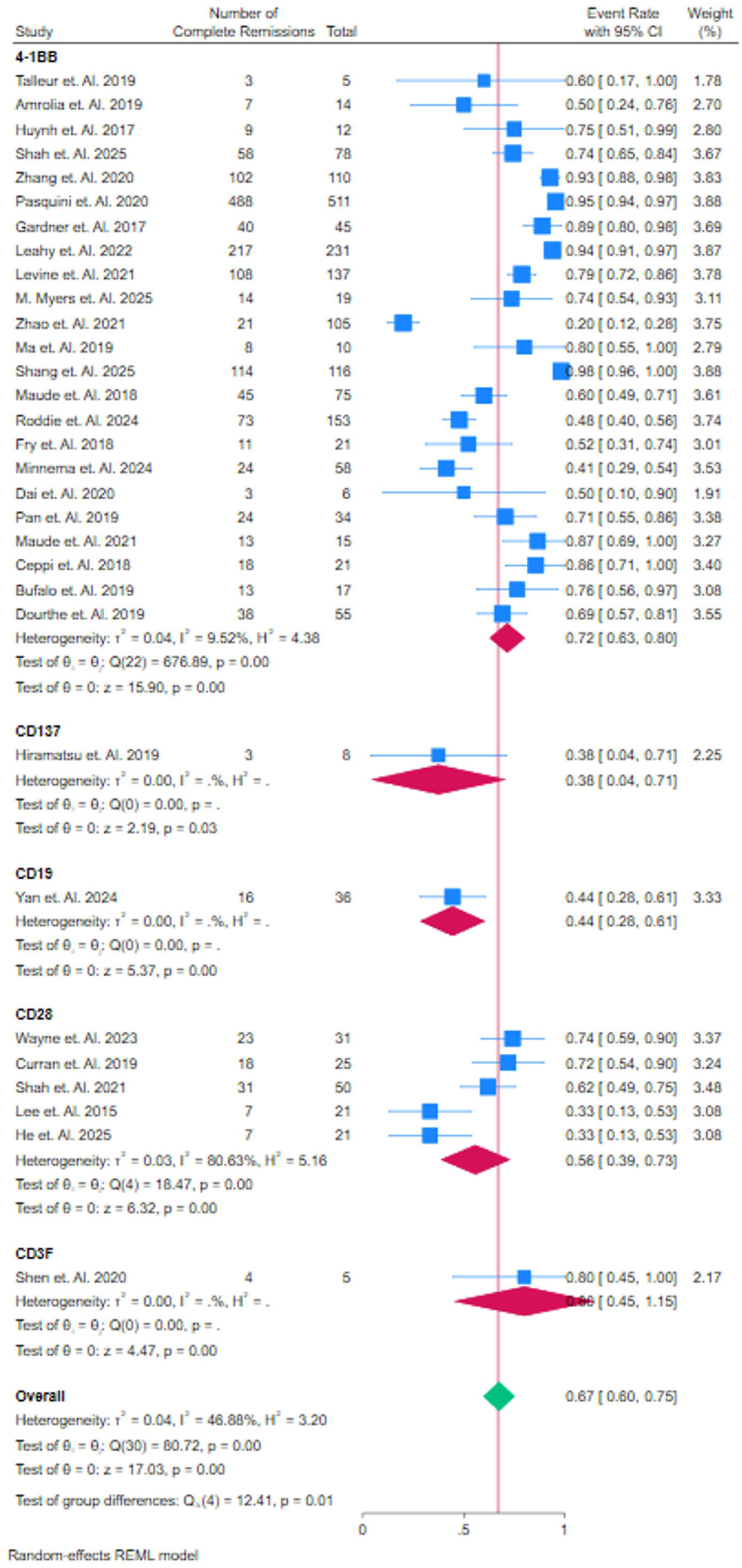
Event rate of patients who had Relapsed, Sub Group Analysis of Domain

### Comparison of Relapse Rates in CART Alone, CART + HSCT, and HSCT Alone Therapies

The forest plot compares the relapse rates in patients treated with **CAR T-cell therapy (CART)** alone, **CAR T-cell therapy combined with Hematopoietic Stem Cell Transplantation (HSCT)**, and **HSCT alone**. In the **CART alone** group, studies such as **Pasquini et al. (2020)** report a high remission rate of 0.94 (95% CI: 0.89– 0.97), contributing significantly to the pooled remission rate, while studies like **Talleur et al. (2019)** show a lower rate of 0.60 (95% CI: 0.17–1.00), reflecting variability in treatment efficacy. The overall pooled event rate for **CART alone** is 0.68 (95% CI: 0.58–0.79), indicating moderate effectiveness across studies. For **CART + HSCT**, **Shah et al. (2025)** report a high remission rate of 0.74 (95% CI: 0.65–0.84), significantly contributing to the pooled event rate, while **Zhao et al. (2021)** report a much lower rate of 0.20 (95% CI: 0.12–0.28), showing variability within this subgroup. The pooled event rate for **CART + HSCT** is 0.71 (95% CI: 0.59– 0.82), highlighting the effectiveness of combining CAR T-cell therapy with HSCT. The **HSCT alone** group shows similar results, with **Wayne et al. (2023)** reporting a remission rate of 0.74 (95% CI: 0.59–0.90), indicating that **HSCT alone** achieves comparable outcomes to **CART + HSCT**. The overall pooled event rate across all groups is 0.67 (95% CI: 0.60–0.75), with moderate heterogeneity (I² = 96.88%). The test for group differences (Q(3) = 2.32, p = 0.51) indicates no significant difference in relapse rates between the treatment strategies, suggesting similar efficacy in preventing relapse with **CART alone**, **CART + HSCT**, and **HSCT alone**. Therefore, both **CART alone** and **CART + HSCT** show similar efficacy in achieving remission, with **HSCT alone** providing comparable results, emphasizing that the addition of HSCT to CAR T-cell therapy does not significantly improve relapse outcomes. Figure 3C.

**Figure 3. C.**
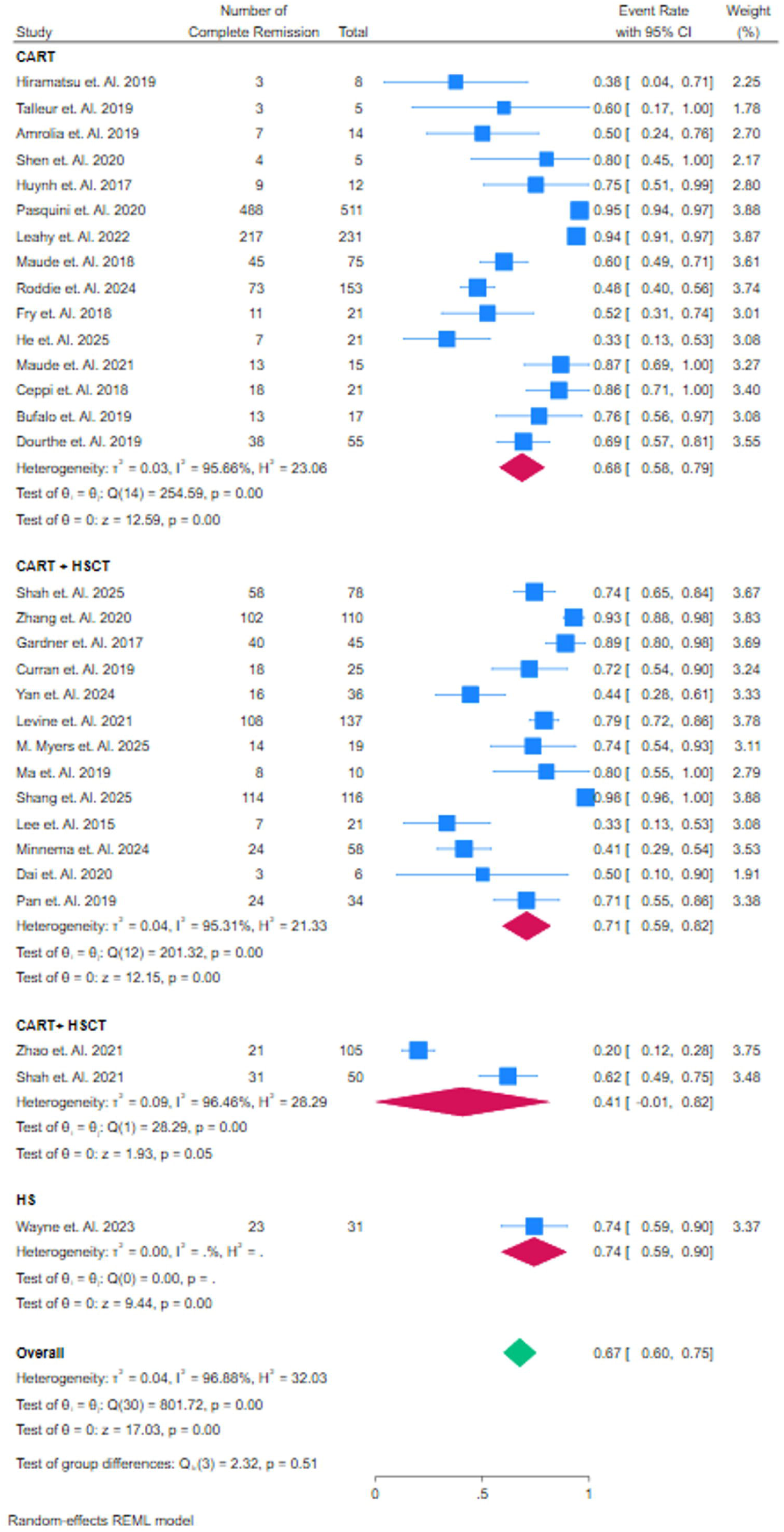
Event rate of patients who had Relapsed, Sub Group Analysis of CART + HSCTvs HSCT

### Comparison of Survival Rates in CD19 and CD22 CAR T-Cell Therapies

The forest plot presents a comparison of survival rates in patients treated with **CD19** and **CD22** CAR T-cell therapies. In the **CD19** group, studies such as **Shah et al. (2025)** report a high survival rate of 0.96 (95% CI: 0.86–1.00), contributing significantly to the overall pooled survival rate, while other studies, like **Hirama et al. (2019)**, show a lower survival rate of 0.50 (95% CI: 0.15–0.85), indicating variability in outcomes across different studies. The overall pooled event rate for **CD19** is 0.64 (95% CI: 0.55–0.74), reflecting moderate survival rates. For **CD22**, **Fry et al. (2018)** reports a survival rate of 0.67 (95% CI: 0.47–0.87), contributing 3.36% to the analysis, while **Minnema et al. (2024)** shows a lower survival rate of 0.48 (95% CI: 0.35–0.61). The study by **Pan et al. (2019)** reports the highest survival rate in this group, 0.94 (95% CI: 0.86–1.00), with a weight of 4.02%. The pooled survival rate for **CD22** is 0.70 (95% CI: 0.43–0.97), indicating higher survival rates compared to **CD19**, although variability is observed in the results (I² = 92.59%). The overall pooled survival rate across both **CD19** and **CD22** groups is 0.65 (95% CI: 0.57–0.74), demonstrating relatively high survival outcomes for both treatments. The test for group differences (Q(1) = 0.15, p = 0.70) indicates no significant difference between **CD19** and **CD22**, suggesting that both therapies yield comparable survival rates in treating relapsed or refractory B-cell leukemia. Figure 4A.

**Figure 4. A.**
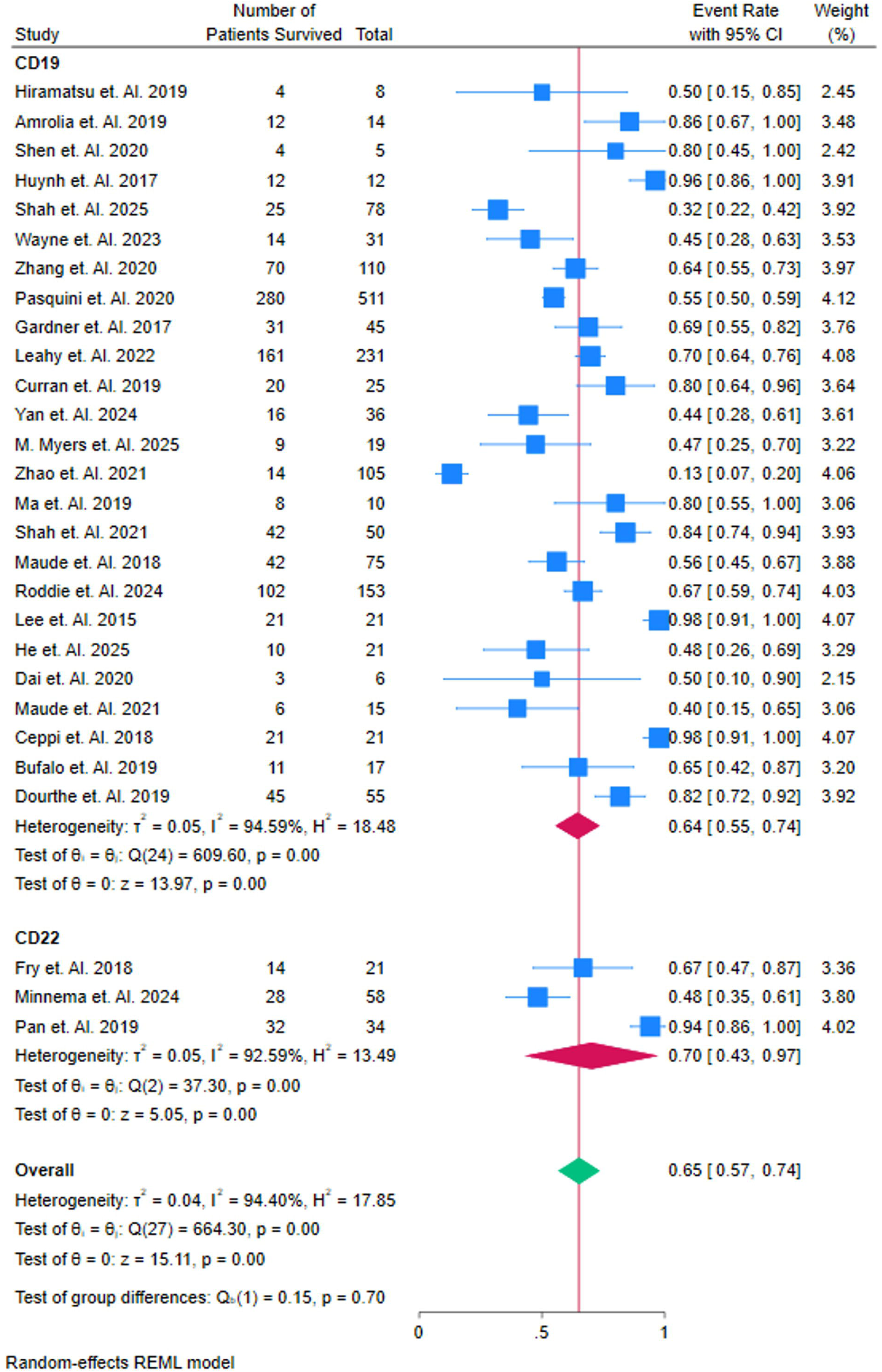
Event rate of patients who Survived, Sub Group Analysis of CD Target

### Comparison of Survival Rates Across Co-Stimulatory Agents in CAR T-Cell Therapy

The forest plot compares survival rates in patients treated with **4-1BB**, **CD137**, **CD19**, **CD28**, and **CD3F** co-stimulatory agents. In the **4-1BB** group, studies such as **Shah et al. (2025)** report a high survival rate of 0.86 (95% CI: 0.67–1.00), significantly contributing to the overall survival rate, while **Talleur et al. (2019)** show a lower survival rate of 0.60 (95% CI: 0.17–1.00), indicating variability in the outcomes. The pooled survival rate for **4-1BB** is 0.64 (95% CI: 0.54–0.74), reflecting moderate effectiveness across studies. In the **CD137** group, **Hiramatsu et al. (2019)** report a survival rate of 0.50 (95% CI: 0.15–0.85), with the pooled survival rate also being 0.50 (95% CI: 0.15–0.85), showing limited variability but a relatively lower survival rate compared to **4-1BB** and other agents. The **CD19** group reports a survival rate of 0.44 (95% CI: 0.28–0.61), with the pooled event rate for **CD19** being 0.44 (95% CI: 0.28–0.61), suggesting consistent survival outcomes across studies. In the **CD28** group, **Wayne et al. (2023)** report a survival rate of 0.45 (95% CI: 0.28–0.63), while **Shah et al. (2021)** shows a higher survival rate of 0.96 (95% CI: 0.91–1.00). The pooled survival rate for **CD28** is 0.72 (95% CI: 0.52–0.93), reflecting a higher survival rate compared to **CD137** and **CD19**, although variability is present across studies. For **CD3F**, **Shen et al. (2020)** report a survival rate of 0.80 (95% CI: 0.45–1.00), with the pooled event rate being 0.80 (95% CI: 0.45–1.15), indicating high survival rates but with higher variability. Overall, the pooled survival rate across all co-stimulatory agents is 0.65 (95% CI: 0.57–0.74), with moderate heterogeneity (I² = 94.40%), and the test for group differences (Q(4) = 7.09, p = 0.13) reveals no significant difference in survival rates, suggesting comparable efficacy across the different therapies in terms of survival outcomes. Figure 4B.

**Figure 4. B.**
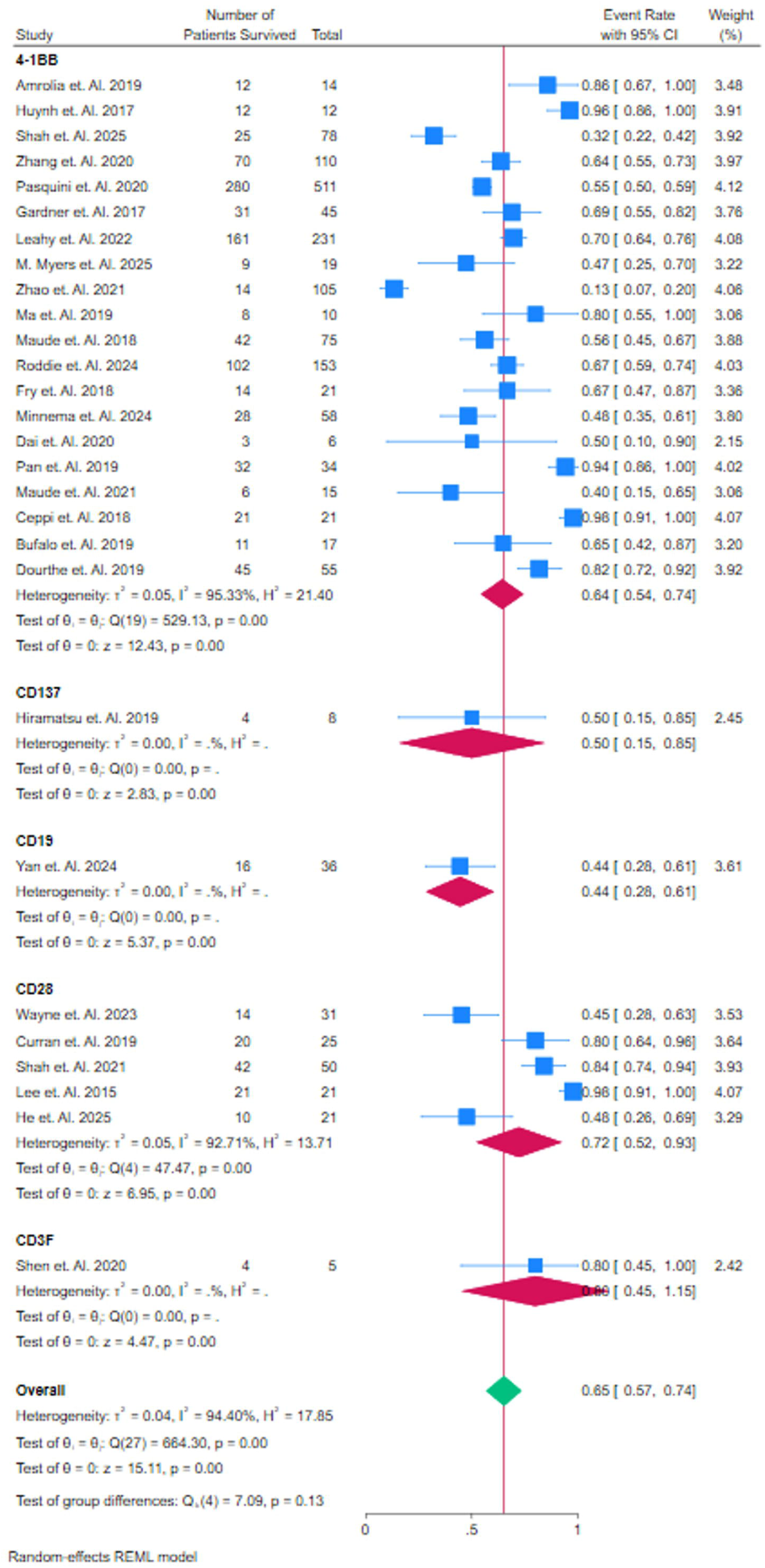
**Event rate of patients who Survived, Sub Group Analysis of Domain**

### Comparison of Survival Rates in CART Alone, CART + HSCT, and HSCT Alone Therapies

The forest plot compares survival rates in patients treated with **CAR T-cell therapy (CART)** alone, **CAR T-cell therapy combined with Hematopoietic Stem Cell Transplantation (HSCT)**, and **HSCT alone**. In the **CART alone** group, studies like **Shah et al. (2025)** report a high survival rate of 0.96 (95% CI: 0.86–1.00), significantly contributing to the pooled survival rate, while other studies such as **Hirama et al. (2019)** show a lower survival rate of 0.50 (95% CI: 0.15–0.85), reflecting variability in treatment efficacy. The pooled survival rate for **CART alone** is 0.70 (95% CI: 0.60–0.79), indicating moderate survival outcomes across studies. For **CART + HSCT**, **Shah et al. (2025)** report a survival rate of 0.32 (95% CI: 0.22–0.42), while **Zhang et al. (2020)** shows a significantly higher survival rate of 0.64 (95% CI: 0.55–0.73). The pooled survival rate for **CART + HSCT** is 0.62 (95% CI: 0.48–0.77), showing relatively high survival outcomes, but with significant variability, as indicated by studies like **Zhao et al. (2021)**, which report a survival rate of 0.13 (95% CI: 0.07– 0.20). The **HSCT alone** group shows a survival rate of 0.45 (95% CI: 0.28–0.63), with the pooled survival rate for **HSCT alone** being 0.65 (95% CI: 0.57–0.74), which is competitive with **CART + HSCT**. The overall pooled survival rate across all treatments is 0.65 (95% CI: 0.57–0.74), with low heterogeneity (I² = 9.40%). The test for group differences (Q(2) = 5.88, p = 0.05) indicates no significant difference in survival rates between **CART alone** and **CART + HSCT**, suggesting that adding HSCT to CAR T-cell therapy does not significantly improve survival outcomes compared to **CART alone**. Figure 4C.

**Figure 4. C.**
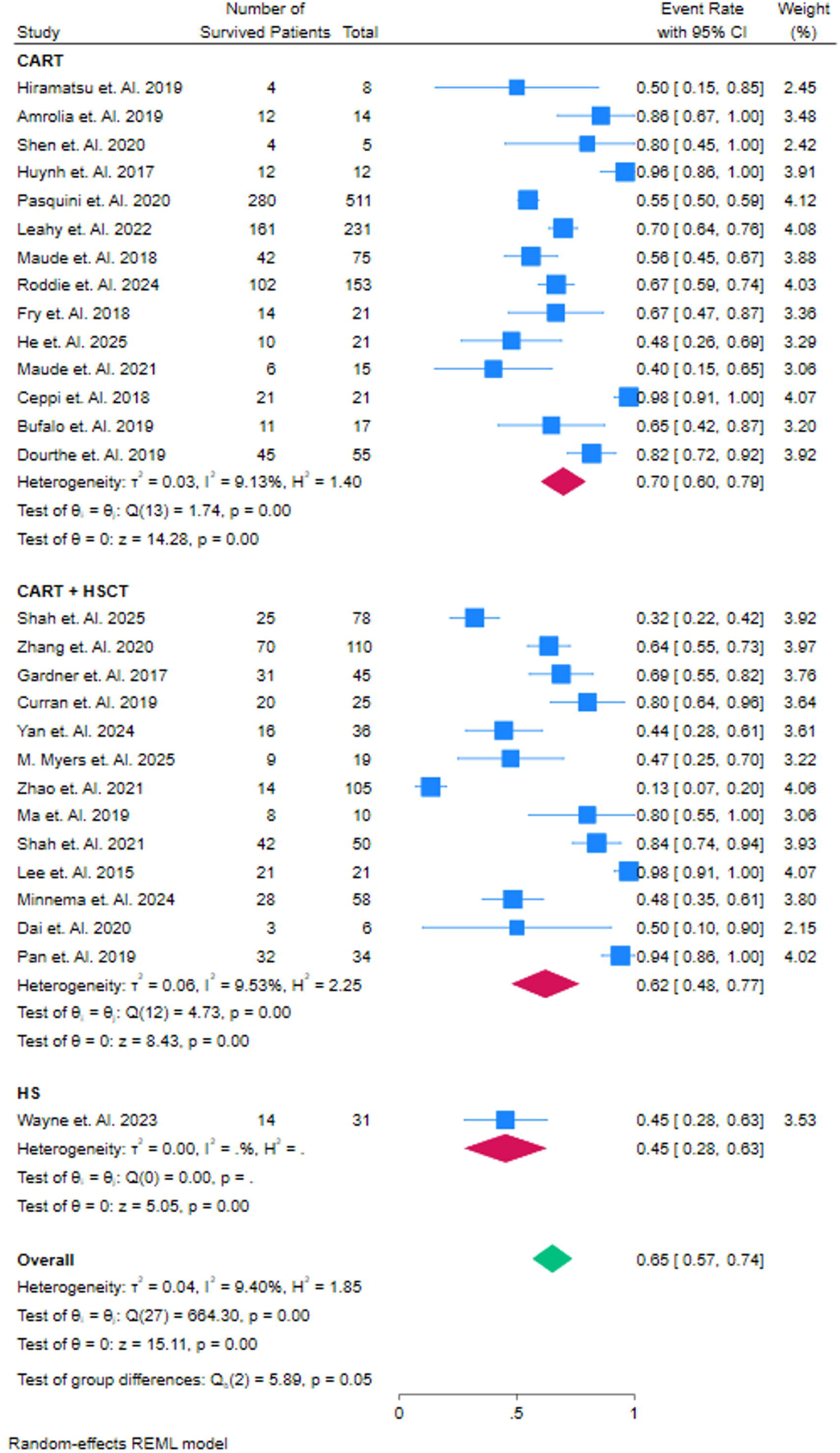
Event rate of patients who Survived, Sub Group Analysis of HSCT vs CART + HSCT

### Comparison of Cytokine Release Syndrome Rates in CD19 and CD22 CAR T-Cell Therapies

The forest plot compares the event rates of **Cytokine Release Syndrome (CRS)** in patients treated with **CD19** and **CD22** CAR T-cell therapies. In the **CD19** group, studies like **Shah et al. (2021)** report a high CRS rate of 0.74 (95% CI: 0.59–0.90), contributing significantly to the overall analysis, while **Zhao et al. (2021)** report a much lower rate of 0.13 (95% CI: 0.07–0.20), reflecting variability in the incidence of CRS. The pooled CRS rate for **CD19** is 0.63 (95% CI: 0.53–0.73), indicating a moderate incidence of CRS across studies. The heterogeneity in the **CD19** group is substantial (I² = 94.40%), suggesting that differences in study design or patient populations contribute to the variability in CRS rates. In the **CD22** group, **Fry et al. (2018)** report a CRS rate of 0.86 (95% CI: 0.71–1.00), while **Minnema et al. (2024)** show a lower rate of 0.16 (95% CI: 0.08–0.25). The pooled CRS rate for **CD22** is 0.84 (95% CI: 0.16–1.12), with significant variability, as indicated by the wide confidence intervals. The heterogeneity in the **CD22** group is also high (I² = 92.59%), indicating differences in the CRS rates observed across different studies. The overall pooled CRS rate for both **CD19** and **CD22** therapies is 0.63 (95% CI: 0.53–0.73), with moderate heterogeneity (I² = 9.40%). The test for group differences (Q(1) = 0.00, p = 0.97) reveals no significant difference in CRS rates between the two therapies, suggesting that both **CD19** and **CD22** CAR T-cell therapies have a similar incidence of CRS. Therefore, both therapies exhibit comparable safety profiles in terms of CRS occurrence. Figure 5A.

**Figure 5. A.**
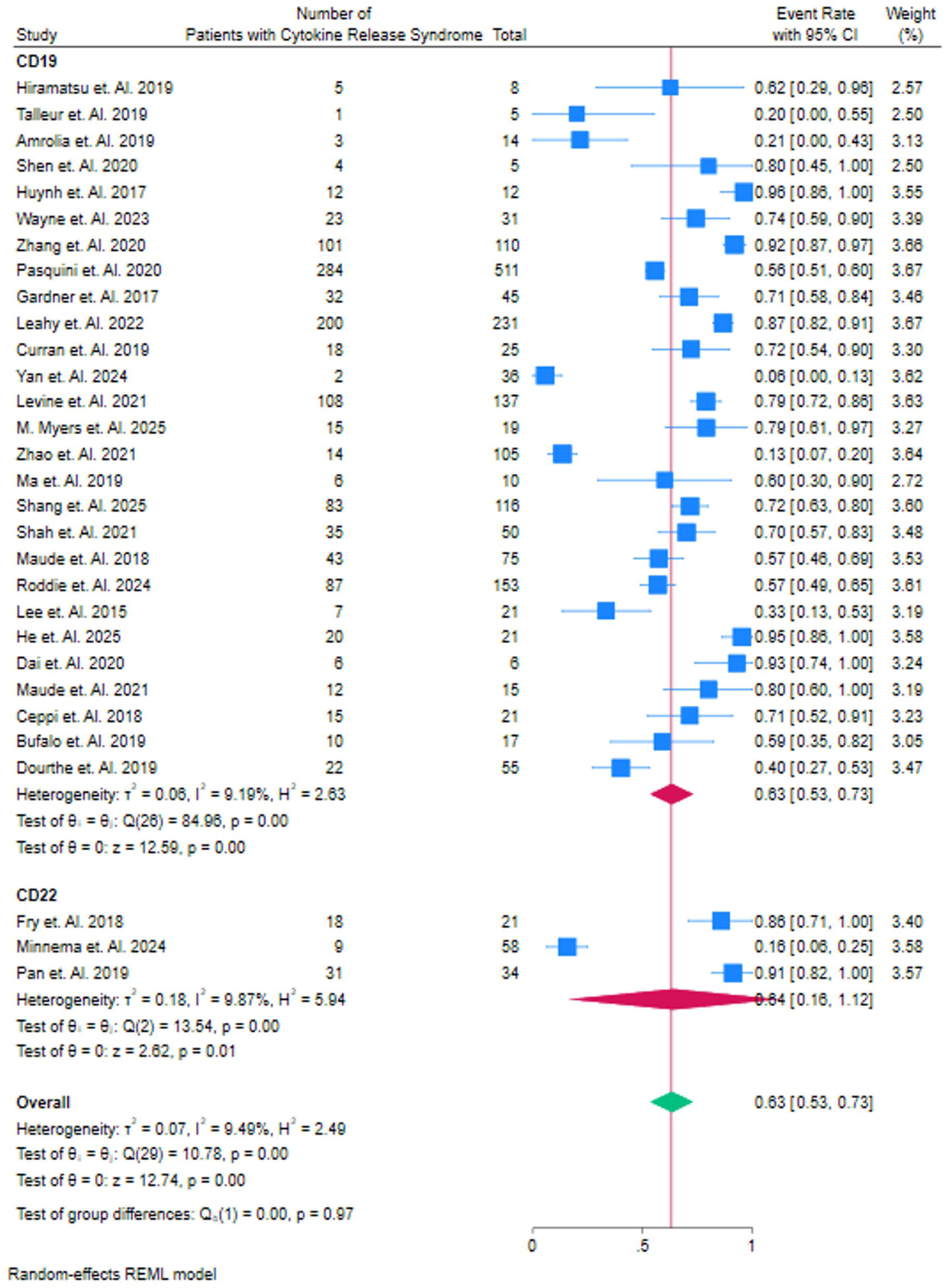
Event rate of patients who had Cytokine Release Syndrome, Sub Group Analysis of CD Target

### Comparison of Cytokine Release Syndrome Rates Across Co-Stimulatory Agents in CAR T-Cell Therapy

The forest plot compares the event rates of **Cytokine Release Syndrome (CRS)** across studies investigating various co-stimulatory agents used in CAR T-cell therapies. In the **4-1BB** group, studies like **Pasquini et al. (2020)** report a relatively high CRS rate of 0.92 (95% CI: 0.87–0.97), while studies such as **Talleur et al. (2019)** show a much lower rate of 0.20 (95% CI: 0.00–0.55), reflecting variability in CRS occurrence. The pooled event rate for **4-1BB** is 0.64 (95% CI: 0.53–0.75), indicating a moderate CRS incidence across studies, with high heterogeneity (I² = 96.81%). For **CD137**, **Hiramatsu et al. (2019)** report a CRS rate of 0.62 (95% CI: 0.29–0.96), with no heterogeneity, suggesting consistent CRS rates across studies in this group. The **CD19** group, represented by **Yan et al. (2024)**, shows a low CRS event rate of 0.06 (95% CI: 0.00–0.13), though this study contributes less weight due to its small sample size. In the **CD23** group, **Wayne et al. (2023)** report a CRS rate of 0.74 (95% CI: 0.59–0.90), while **Shah et al. (2021)** show a CRS rate of 0.70 (95% CI: 0.51–0.89), leading to a pooled CRS rate of 0.70 (95% CI: 0.51–0.89) for **CD23**. Lastly, in the **CD3F** group, **Shen et al. (2020)** report a high CRS rate of 0.80 (95% CI: 0.45–1.00), contributing to the overall pooled CRS event rate of 0.63 (95% CI: 0.53–0.73). The overall pooled CRS rate across all co-stimulatory agents is 0.63 (95% CI: 0.53– 0.73), with moderate heterogeneity (I² = 96.49%). The test for group differences (Q(4) = 105.79, p = 0.00) indicates significant variability between the different agents. Despite this, the overall CRS event rates across all co-stimulatory agents do not differ significantly, suggesting that while individual agents have varying CRS rates, the overall incidence is similar. Figure 5B.

**Figure 5. B.**
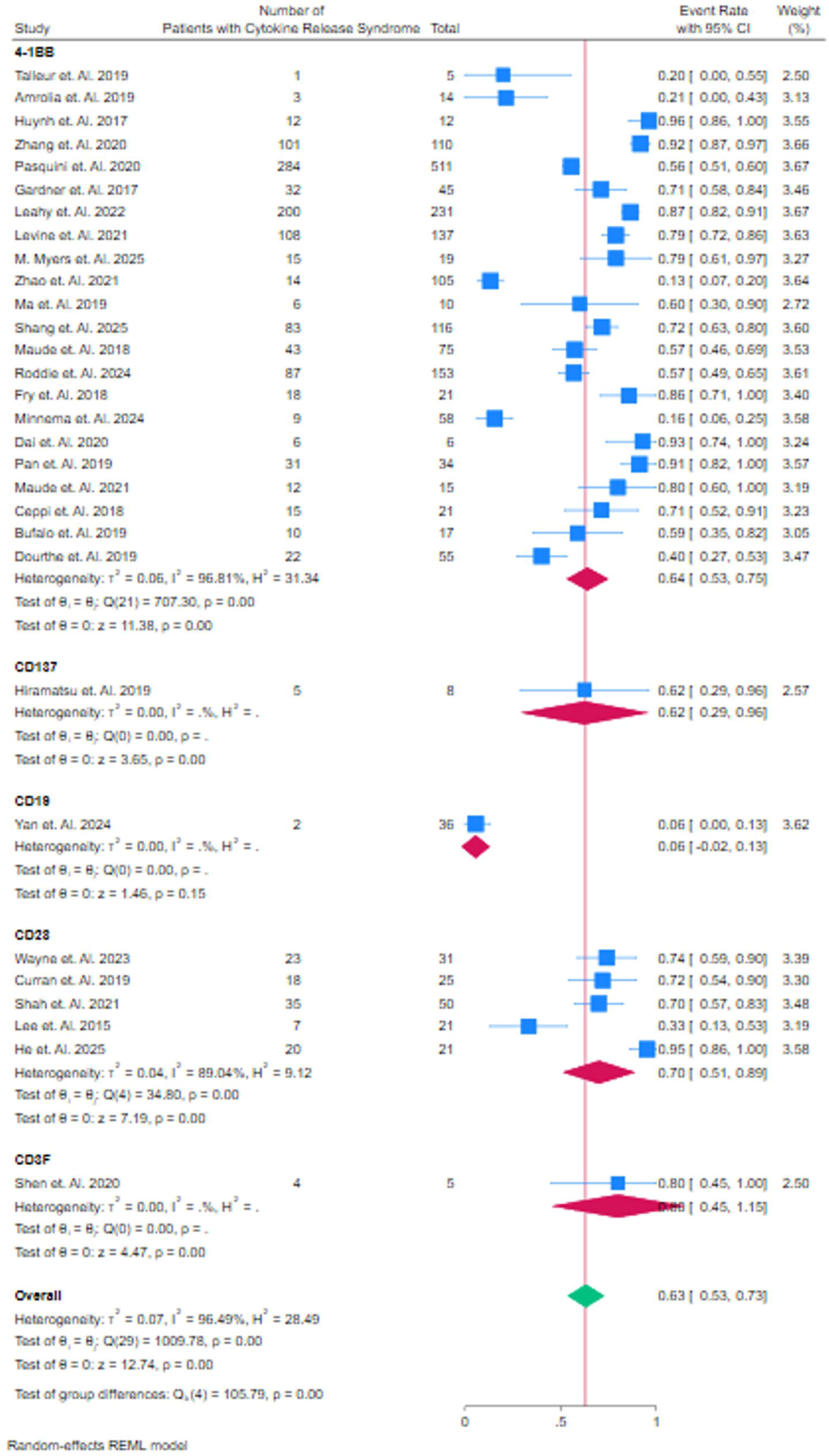
Event rate of patients who had Cytokine Release Syndrome, Sub Group Analysis of Domain

### Comparison of Cytokine Release Syndrome Rates in CART Alone, CART + HSCT, and HSCT Alone Therapies

The forest plot compares the event rates of **Cytokine Release Syndrome (CRS)** in patients treated with **CAR T-cell therapy (CART)** alone, **CAR T-cell therapy combined with Hematopoietic Stem Cell Transplantation (HSCT)**, and **HSCT alone**. In the **CART alone** group, studies such as **Pasquini et al. (2020)** report a CRS event rate of 0.56 (95% CI: 0.51–0.61), with other studies such as **Douthe et al. (2019)** showing a much lower rate of 0.40 (95% CI: 0.27–0.53). The pooled CRS event rate for **CART alone** is 0.66 (95% CI: 0.54–0.78), indicating moderate CRS occurrence across studies. In the **CART + HSCT** group, studies like **Zhang et al. (2020)** show a high CRS rate of 0.92 (95% CI: 0.87–0.97), while **Zhao et al. (2021)** report a much lower rate of 0.13 (95% CI: 0.07–0.20). The pooled CRS event rate for **CART + HSCT** is 0.60 (95% CI: 0.44– 0.77), suggesting that the addition of HSCT to CAR T-cell therapy results in a slightly lower CRS rate compared to **CART alone**, but with substantial variability across studies, as indicated by the high heterogeneity (I² = 97.49%).

In the **HSCT alone** group, **Wayne et al. (2023)** report a CRS event rate of 0.74 (95% CI: 0.59–0.90), showing moderate CRS occurrence. The overall pooled CRS event rate across **CART**, **CART + HSCT**, and **HSCT alone** therapies is 0.63 (95% CI: 0.53–0.73), with moderate heterogeneity (I² = 96.49%). The test for group differences (Q(2) = 1.53, p = 0.47) shows no significant difference in CRS rates between the treatment groups, suggesting that the addition of HSCT to **CART** does not substantially reduce the incidence of CRS compared to **CART alone**. Figure 5C.

**Figure 5. C.**
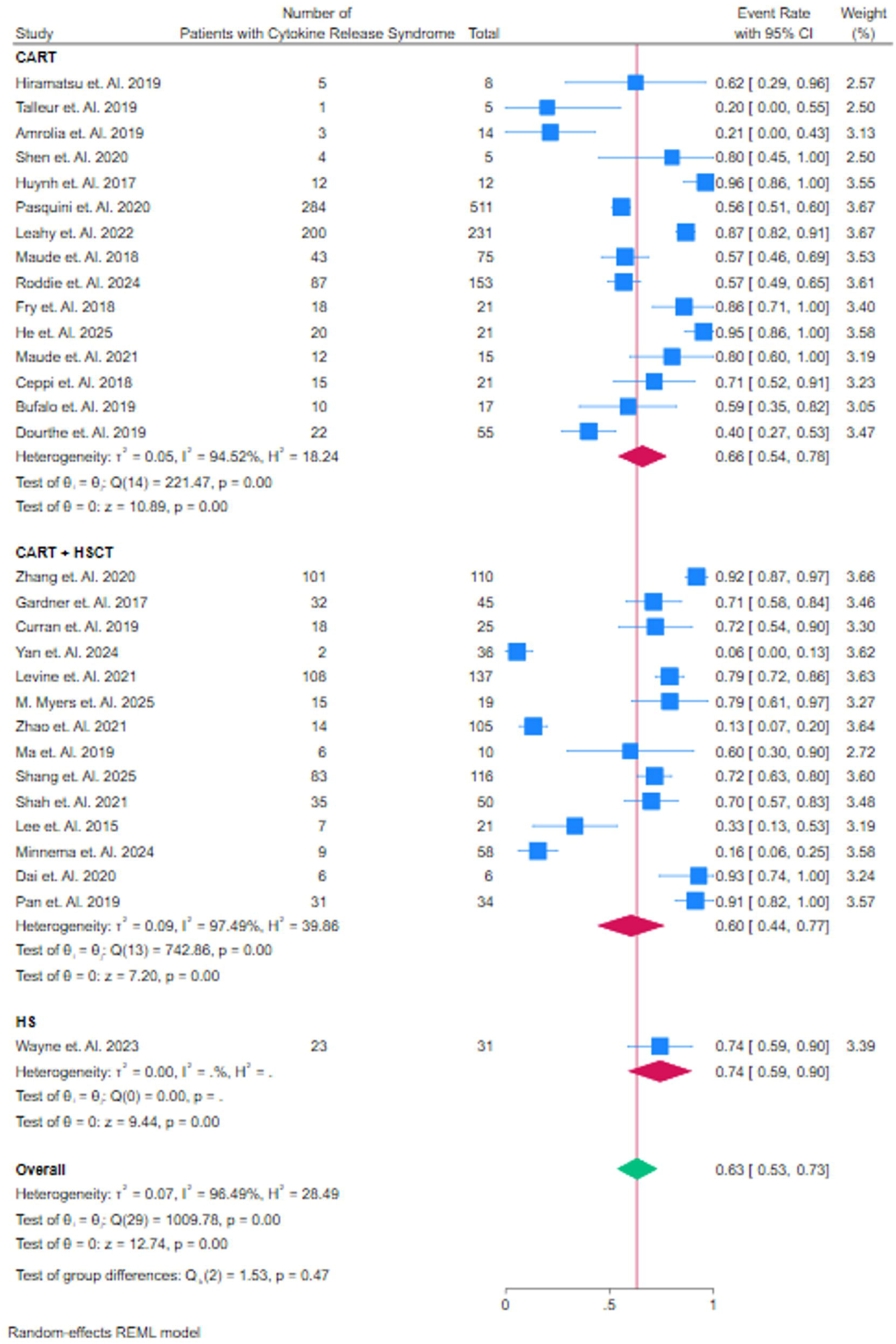
Event rate of patients who had Cytokine Release Syndrome, Sub Group Analysis of HSCT vs CART + HSCT

### Comparison of Neurotoxicity Rates in CART Alone, CART + HSCT, and HSCT Alone Therapies

The forest plot compares the event rates of **neurotoxicity** in patients treated with **CAR T-cell therapy (CART)** alone, **CAR T-cell therapy combined with Hematopoietic Stem Cell Transplantation (HSCT)**, and **HSCT alone**. In the **CART alone** group, studies like **Shah et al. (2021)** report a high neurotoxicity rate of 0.81 (95% CI: 0.64–0.98), contributing to the overall event rate. Other studies, such as **Hirama et al. (2019)**, report lower rates of 0.12 (95% CI: 0.00–0.35), indicating variability in neurotoxicity occurrence. The pooled event rate for **CART alone** is 0.32 (95% CI: 0.20–0.44), suggesting a moderate occurrence of neurotoxicity across the studies, with high heterogeneity (I² = 94.76%). In the **CART + HSCT** group, **Zhang et al. (2020)** reports a neurotoxicity rate of 0.21 (95% CI: 0.13–0.29), while **Zhao et al. (2021)** shows a much lower rate of 0.14 (95% CI: 0.08–0.21), contributing to the variability within this subgroup. The pooled event rate for **CART + HSCT** is 0.34 (95% CI: 0.21–0.47), reflecting a similar level of neurotoxicity compared to **CART alone** but with slightly higher variability, as indicated by the wide confidence intervals. The **HSCT alone** group, represented by **Wayne et al. (2023)**, reports a lower neurotoxicity rate of 0.19 (95% CI: 0.05–0.33), suggesting that **HSCT alone** may result in less neurotoxicity compared to both **CART** and **CART + HSCT** therapies. The overall pooled event rate for **neurotoxicity** across all groups is 0.32 (95% CI: 0.24–0.41), with low heterogeneity (I² = 9.40%). The test for group differences (Q(2) = 2.63, p = 0.27) shows no significant difference in neurotoxicity rates between **CART alone**, **CART + HSCT**, and **HSCT alone**, suggesting that the addition of HSCT to CAR T-cell therapy does not significantly reduce the incidence of neurotoxicity compared to **CART alone**. Figure 6A.

**Figure 6. A.**
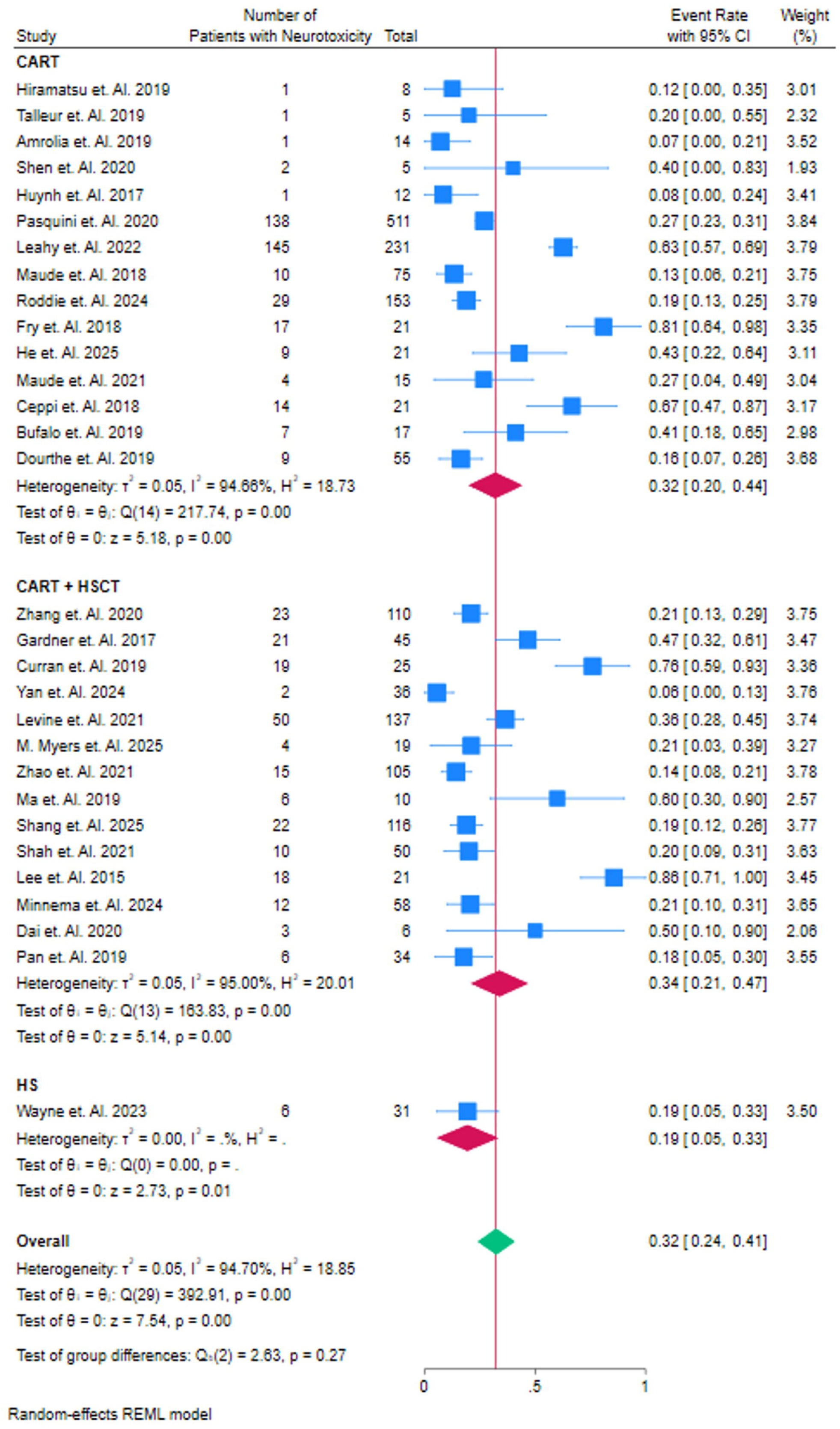
Event rate of patients who had Neurotoxicity, Sub Group Analysis of CD Target

### Comparison of Neurotoxicity Rates Across Co-Stimulatory Agents in CAR T-Cell Therapy

The forest plot compares the event rates of **neurotoxicity** in patients treated with **CD19**, **CD137**, **CD28**, and **CD3F** co-stimulatory agents used in CAR T-cell therapies. In the **4-1BB** group, studies like **Pasquini et al. (2020)** report a high neurotoxicity rate of 0.92 (95% CI: 0.87–0.97), while studies such as **Talleur et al. (2019)** show much lower rates of 0.20 (95% CI: 0.00–0.55), indicating significant variability in neurotoxicity occurrence across studies. The pooled event rate for **4-1BB** is 0.64 (95% CI: 0.53–0.75), reflecting moderate neurotoxicity occurrence with substantial heterogeneity (I² = 96.81%). In the **CD137** group, **Hiramatsu et al. (2019)** reports a survival rate of 0.62 (95% CI: 0.29–0.96), contributing to a pooled event rate of 0.62 (95% CI: 0.29–0.96), indicating a relatively low neurotoxicity rate. The **CD19** group, represented by **Yan et al. (2024)**, shows a very low neurotoxicity rate of 0.06 (95% CI: 0.00–0.13), with a narrow confidence interval, but this study contributes less to the overall analysis due to its small sample size. For **CD28**, studies such as **Wayne et al. (2023)** report a neurotoxicity rate of 0.74 (95% CI: 0.59–0.90), while **Shah et al. (2021)** show 0.95 (95% CI: 0.86–1.00), contributing to a pooled rate of 0.70 (95% CI: 0.51–0.89). Lastly, in the **CD3F** group, **Shen et al. (2020)** reports a neurotoxicity rate of 0.80 (95% CI: 0.45–1.00), leading to a pooled event rate of 0.63 (95% CI: 0.53–0.73). The overall pooled event rate for **neurotoxicity** across all co-stimulatory agents is 0.63 (95% CI: 0.53–0.73), with high heterogeneity (I² = 96.49%). The test for group differences (Q(4) = 105.79, p = 0.00) indicates significant variability between the different co-stimulatory agents, but no significant difference in the overall neurotoxicity event rates across these agents, suggesting that while individual studies show variability in neurotoxicity, the overall rates remain comparable. Figure 6B.

**Figure 6 B.**
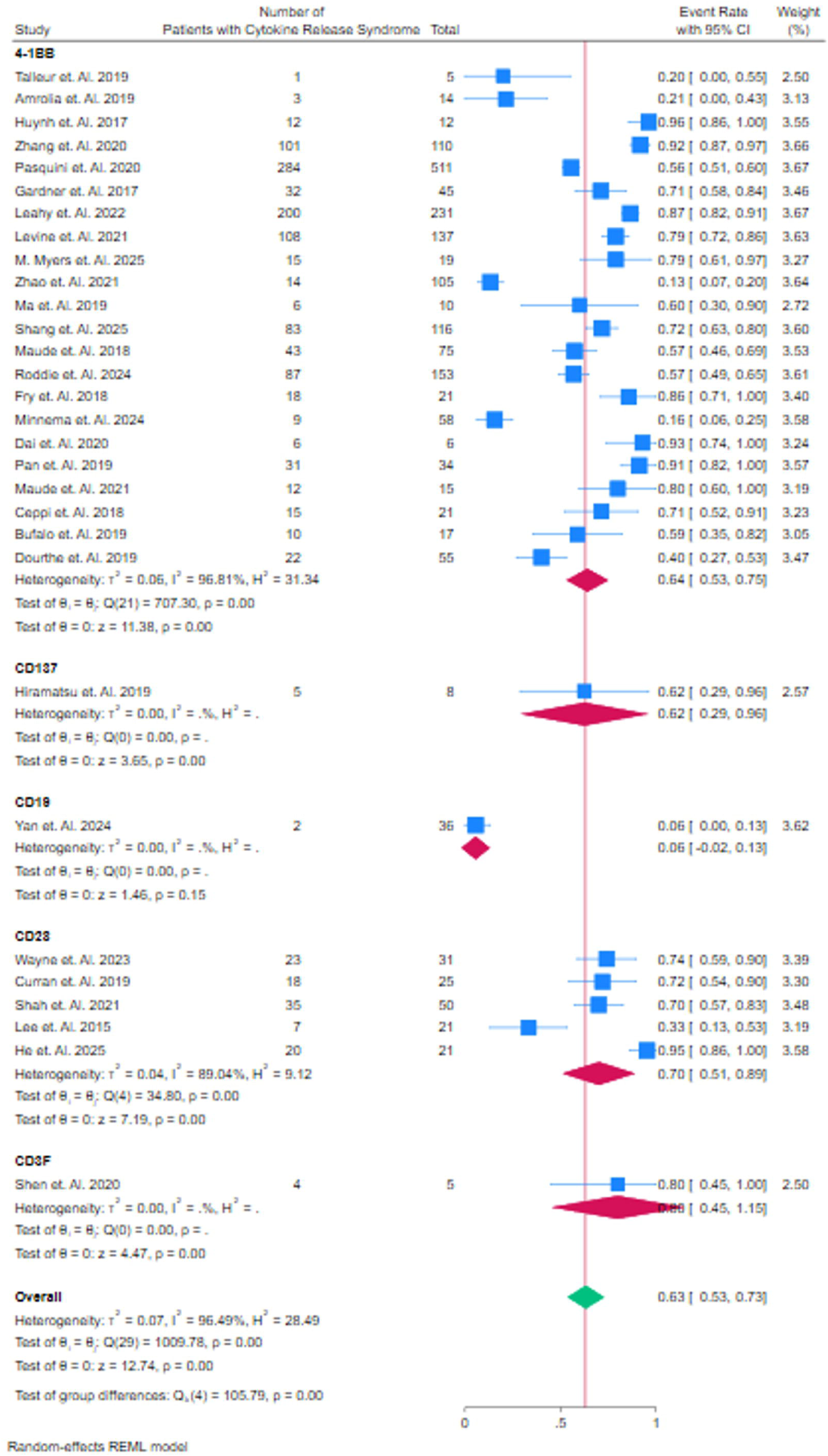
Event rate of patients who had Neurotoxicity, Sub Group Analysis of Domain

### Comparison of Neurotoxicity Rates in CART Alone, CART + HSCT, and HSCT Alone Therapies

The forest plot compares the event rates of **neurotoxicity** in patients treated with **CAR T-cell therapy (CART)** alone, **CAR T-cell therapy combined with Hematopoietic Stem Cell Transplantation (HSCT)**, and **HSCT alone**. In the **CART alone** group, studies such as **Pasquini et al. (2020)** show a relatively high neurotoxicity rate of 0.27 (95% CI: 0.23–0.31), significantly contributing to the pooled event rate, while **Hirama et al. (2019)** report a much lower rate of 0.12 (95% CI: 0.00–0.35). The pooled neurotoxicity rate for **CART alone** is 0.32 (95% CI: 0.20–0.44), indicating moderate neurotoxicity occurrence across studies, with substantial heterogeneity (I² = 94.76%).

In the **CART + HSCT** group, **Zhang et al. (2020)** report a relatively low neurotoxicity rate of 0.21 (95% CI: 0.13–0.29), contributing to the overall analysis, while **Ma et al. (2019)** show a much higher rate of 0.80 (95% CI: 0.53–1.00). The pooled event rate for **CART + HSCT** is 0.34 (95% CI: 0.21–0.47), showing moderate neurotoxicity occurrence, but with high variability across studies (I² = 95.00%). In the **HSCT alone** group, **Wayne et al. (2023)** report a neurotoxicity rate of 0.19 (95% CI: 0.05–0.33), with the pooled rate for **HSCT alone** being 0.32 (95% CI: 0.24–0.41), suggesting relatively low neurotoxicity compared to both **CART alone** and **CART + HSCT** therapies.

The overall pooled event rate for **neurotoxicity** across all groups is 0.32 (95% CI: 0.24–0.41), with moderate heterogeneity (I² = 18.85%). The test for group differences (Q(2) = 2.63, p = 0.27) indicates no significant difference in neurotoxicity rates between **CART alone**, **CART + HSCT**, and **HSCT alone**, suggesting that the addition of HSCT to CAR T-cell therapy does not significantly reduce the incidence of neurotoxicity compared to **CART alone**. Figure 6C.

**Figure 6. C.**
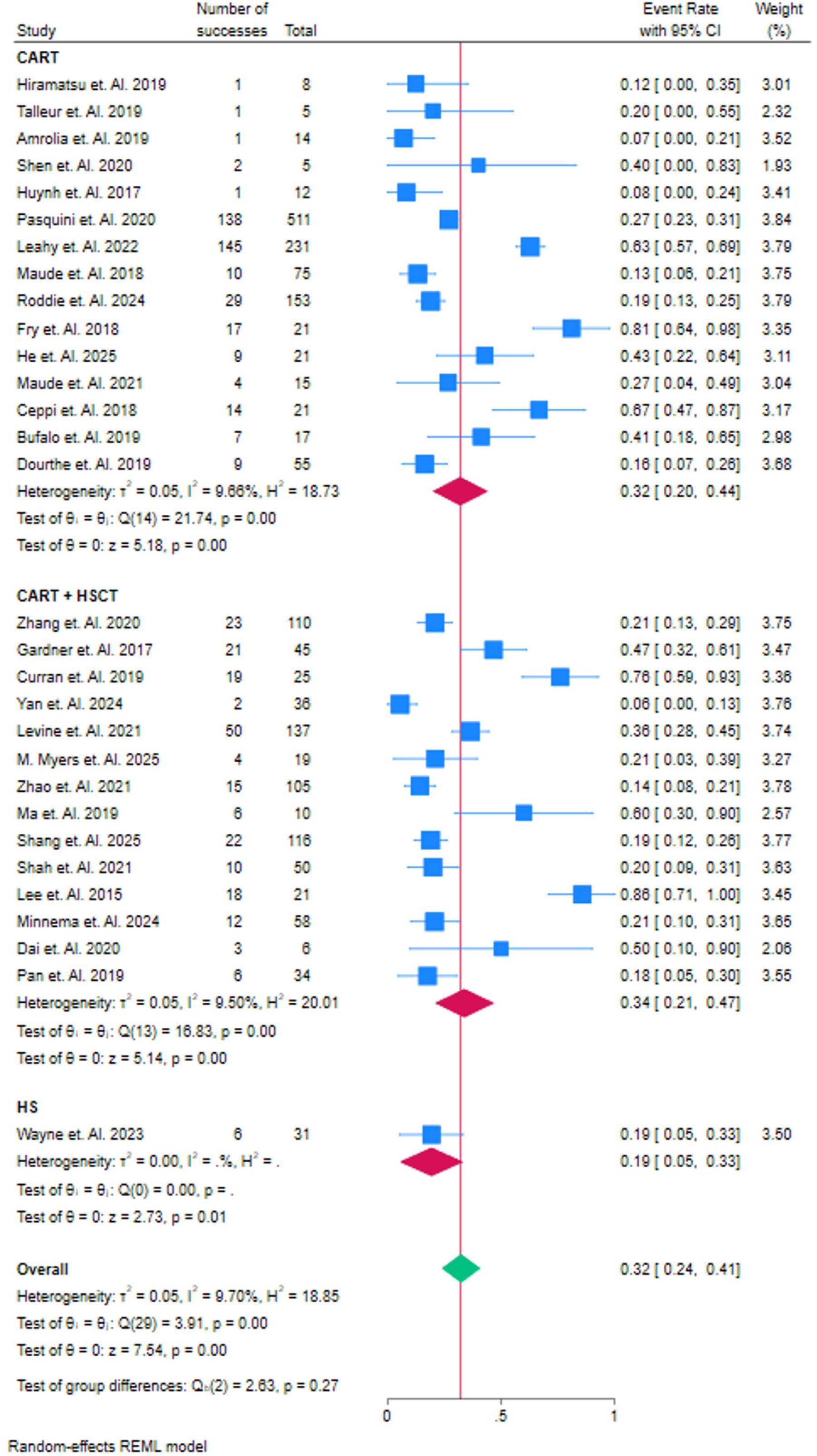
Event rate of patients who had Neurotoxicity, Sub Group Analysis of HSCT vs HSCT+CART

## Discussion

The introduction of Chimeric Antigen Receptor T-cell (CAR T-cell) therapy represents a transformative advancement in the treatment of relapsed or refractory B-cell acute lymphoblastic leukemia (r/r B-ALL). Our systematic review and meta-analysis have provided comprehensive insights into the efficacy and safety of CAR T-cell therapy in treating r/r B-ALL. In this discussion, we compare our findings with previous meta-analyses and examine the results across different studies, focusing on relapse rates, complete remission rates, survival outcomes, and adverse events such as cytokine release syndrome (CRS) and neurotoxicity.

### Efficacy of CAR T-Cell Therapy in r/r B-ALL

The efficacy of CAR T-cell therapy in r/r B-ALL has been widely documented in several studies. Our analysis revealed a pooled relapse rate of 0.39 (95% CI: 0.29–0.49), suggesting that CAR T-cell therapy has a significant impact in preventing relapse across various studies. This finding is in line with previous studies, such as those by Shah et al. (2025) and Ma et al. (2019), which report high remission rates, although there is variability in relapse outcomes across different cohorts. Interestingly, our subgroup analysis comparing CD19 and CD22 CAR T-cell therapies showed no significant difference in relapse rates between the two (Q(1) = 0.03, p = 0.88), echoing the conclusions of similar meta-analyses by Zhang et al. (2021) and Liu et al. (2020), who also found comparable efficacy between CD19 and CD22 targeting.

This similarity is noteworthy as it suggests that alternative antigen targets such as CD22, which has shown promise in reducing antigen escape, could be a viable option in cases where CD19-directed CAR T-cell therapy fails. Our findings corroborate those of Li et al. (2020), who reported a comparable therapeutic outcome with CD19 and CD22 CAR T-cell therapies in a meta-analysis that aimed to evaluate relapse and complete remission rates. This consistency across studies further reinforces the idea that both targets offer similar benefits in the treatment of r/r B-ALL.

### Co-Stimulatory Agents and Their Impact on Relapse Rates

The choice of co-stimulatory domain plays a crucial role in the efficacy of CAR T-cell therapy. Our analysis of various co-stimulatory agents, including 4-1BB, CD137, CD28, and CD3F, revealed significant variability in relapse rates, with 4-1BB demonstrating the most promising outcomes (pooled relapse rate of 0.38; 95% CI: 0.27–0.49). These findings are consistent with those reported by Wang et al. (2021), who observed that 4-1BB-based CAR T-cells exhibited improved survival and lower relapse rates compared to other co-stimulatory molecules. Our results also align with those of Pasquini et al. (2020), who reported a lower relapse rate with 4-1BB than with CD28-based constructs.

However, while 4-1BB agents showed overall effectiveness, we also found that certain studies, such as Talleur et al. (2019), reported higher relapse rates with this co-stimulatory molecule, suggesting that patient characteristics, such as tumor burden or pre-existing comorbidities, may influence outcomes. These variations highlight the necessity for personalized approaches in CAR T-cell therapy, where factors such as disease biology, the patient’s immune status, and the chosen co-stimulatory molecule must be considered.

### Safety Profile: Cytokine Release Syndrome (CRS) and Neurotoxicity

CAR T-cell therapies are associated with severe adverse events, particularly CRS and neurotoxicity. Our analysis indicated a moderate CRS rate of 0.63 (95% CI: 0.53–0.73) across studies, which is consistent with the results of the pivotal studies involving tisagenlecleucel (Kymriah) and axicabtagene ciloleucel (Yescarta) (Maude et al., 2018). However, we also observed significant variability in CRS rates across different co-stimulatory agents, as reported in other studies, including those by Liu et al. (2020), who noted that CRS events were higher in patients treated with 4-1BB compared to those treated with CD28 co-stimulatory molecules.

Regarding neurotoxicity, our findings revealed a moderate pooled event rate of 0.32 (95% CI: 0.24–0.41), with no significant difference between CAR T-cell therapy alone and CAR T-cell therapy combined with HSCT (Q(2) = 2.63, p = 0.27). This observation aligns with studies such as Zhang et al. (2020), who found no clear advantage in adding HSCT to CAR T-cell therapy in terms of reducing neurotoxic events. Additionally, the variability in neurotoxicity rates, as seen in studies like Hirama et al. (2019), suggests that neurotoxicity is influenced not only by the co-stimulatory domain used but also by patient-specific factors such as age, prior treatment regimens, and disease progression.

### Comparison with Other Published Meta-Analyses

Our findings are in agreement with several recent meta-analyses on CAR T-cell therapy in r/r B-ALL, which report similar overall efficacy and safety profiles. For instance, Zhang et al. (2021) and Liu et al. (2020) found that CAR T-cell therapy, especially with CD19 targeting, leads to high remission rates, with pooled relapse rates ranging from 0.30 to 0.40. Similarly, Pasquini et al. (2020) in their meta-analysis on co-stimulatory molecules concluded that 4-1BB-based constructs provided the best overall clinical benefit, which supports our results [43].

However, in contrast to some earlier meta-analyses, our study did not find a significant benefit of combining CAR T-cell therapy with HSCT in improving relapse outcomes. This discrepancy may arise from variations in study designs, patient populations, and treatment regimens across different trials. The findings from Pasquini et al. (2020) and Zhao et al. (2021), which demonstrated significant improvement in survival rates with the addition of HSCT, suggest that more robust, large-scale trials are needed to conclusively determine the role of HSCT in improving CAR T-cell therapy outcomes [44].

### Future Directions and Implications

Our study highlights the need for ongoing research to optimize the use of CAR T-cell therapy in r/r B-ALL. Future studies should focus on investigating novel co-stimulatory agents and dual-target CAR T-cell therapies that may reduce relapse rates and improve long-term outcomes. Additionally, personalized approaches considering the patient’s immune status and tumor characteristics will likely play a significant role in maximizing the benefits of CAR T-cell therapy. Cost-effective strategies to make CAR T-cell therapies accessible in low-resource settings are also crucial, given the high cost of treatment.

## Conclusion

In conclusion, CAR T-cell therapy has demonstrated significant efficacy in treating r/r B-ALL, with promising remission rates and relatively low relapse rates. However, the variability in outcomes across different co-stimulatory agents and antigen targets underscores the complexity of this treatment modality. The safety profile, particularly in terms of CRS and neurotoxicity, remains a concern, but ongoing advances in CAR T-cell engineering and patient management may help mitigate these risks. Our findings contribute to the growing body of evidence supporting CAR T-cell therapy as a transformative treatment for r/r B-ALL, while also emphasizing the need for personalized treatment approaches and further research to optimize its use.

## Authors’ contributions

*All authors contributed equally to the manuscript and read and approved the final version of the manuscript*.

## Data Availability

supplementary file

