## supplementary file for "Efficacy and Safety of CAR T-cell Therapy in Relapsed/Refractory B-cell Acute Lymphoblastic Leukemia: A Systematic Review and Meta-analysis"

Supplementary Files

Search String

("CAR T-cell therapy" OR "chimeric antigen receptor T-cell therapy" OR "CAR-T" OR "CAR-T cells")

AND

("relapsed" OR "refractory" OR "relapsed/refractory")

AND

("B-Cell Acute Lymphoblastic Leukemia" OR "B-ALL" OR "acute lymphoblastic leukemia" OR "ALL")

AND

("efficacy" OR "effectiveness" OR "clinical outcomes" OR "response rate" OR "remission rate")

AND

("safety" OR "adverse events" OR "toxicity" OR "cytokine release syndrome" OR "neurotoxicity" OR "ICANS")

AND

("systematic review" OR "meta-analysis" OR "review" OR "pooled analysis")

Table S1. Demographics and Summary of Findings

| **Ref Number** | **Author Name Year** | **Country** | **Total Sample** | **Male** | **Female** | **Rx Name** | **Rx number** | **Cx Name** | **Cx Number** | **Study Design** | **Follow up in months** | **Mean Age** | **Car T-Cell Intervention Type** | **Dosage x 10 ^6** | **CD Target** | **Co-stimulatory Domain** | **CART + HSCT / HSCT** | **GRADE** | **Main Finding** |
| --- | --- | --- | --- | --- | --- | --- | --- | --- | --- | --- | --- | --- | --- | --- | --- | --- | --- | --- | --- |
| 12 | Hiramatsu et. Al. 2019 | Japan | 8 | 5 | 3 | Tisagenlecleucel | 8 | N/A | N/A | Single Arm | 60 | 16 | Tisagenlecleucel | 0.2-0.5 | CD19 | CD137 | CART | High | Tisagenlecleucel (CAR T-cell therapy) demonstrated high remission rates and durable responses in Japanese pediatric and young adult patients with relapsed/refractory B-cell acute lymphoblastic leukemia (r/r B-ALL). Although adverse events such as cytokine release syndrome (CRS) were common, they were manageable |
| 13 | Talleur et. Al. 2019 | USA | 5 | 3 | 2 | Autologous CD19-CAR T-cells | 5 | N/A | N/A | Single Arm | 1 | 21 | Autologous CD19-CAR T-cells | 1 | CD19 | **4-1BB** | CART | High | The study's main finding suggests **encouraging safety, consistent CAR T-cell expansion, and anti-ALL activity** in patients treated with CD19-CAR T-cells, though more data from the ongoing expansion cohort is needed. |
| 14 | Amrolia et. Al. 2019 | UK | 14 | 13 | 1 | CAR T-Cells | 14 | N/A | N/A | Single Arm | 24 | 24 | CAR T-Cells | 1 | CD19 | 4-1BB | CART | High | The study demonstrated **enhanced CAR T-cell expansion and persistence**, leading to **improved disease control** in pediatric ALL patients . |
| 15 | Shen et. Al. 2020 | China | 5 | 2 | 3 | CD19 CART | 5 | N/A | N/A | Observational Cohort | 4 | 12 | CAR T-Cells | 1 | CD19 | CD3F | CART | High | The study demonstrates that **CD19CAR-T cell therapy** can be administered safely under real-time monitoring of Th1/Th2 cytokine patterns in children with relapsed and refractory B-ALL, although side effects like cytokine release syndrome (CRS) and neurotoxicity were observed |
| 16 | Huynh et. Al. 2017 | USA | 12 | 8 | 4 | KTE-C19 | 11 | N/A | N/A | Single Arm | 3.8 | 12 | CAR T-Cells | 2 | CD19 | 4-1BB | CART | High | KTE-C19 demonstrated promising efficacy with a manageable safety profile, producing deep remissions in heavily pre-treated patients with high disease burden |
| 17 | Shah et. Al. 2025 | USA | 78 | 39 | 39 | Brexucabtagene Autoleucel | 78 | N/A | N/A | Single Arm | 41.6 | 42.5 | CAR T-Cells | 1 | CD19 | 4-1BB | CART + HSCT | High | The main finding was that **Brexucabtagene Autoleucel (brexu-cel)** provides long-term benefits for adult patients with relapsed/refractory B-cell acute lymphoblastic leukemia (B-ALL), with a median overall survival of **25.6 months** and durable responses observed even without subsequent HSCT . |
| 18 | Wayne et. Al. 2023 | USA | 31 | 15 | 9 | KTE-X19 | 24 | N/A | N/A | Single Arm | 36.1 | 13.5 | CAR T-Cells | 2 | CD19 | CD28 | HS | High | KTE-X19 treatment achieved a **67% overall remission rate** and a **100% MRD-negativity rate** in responders, with many patients proceeding to **alloSCT** |
| 19 | Zhang et. Al. 2020 | China | 110 | 68 | 42 | CAR T-Cells | 110 | N/A | N/A | Single Arm | 7.8 | 12 | CAR T-Cells | 1 | CD19 | 4-1BB | CART + HSCT | High | Anti-CD19 CAR T-cell therapy is **safe and effective** in patients with relapsed/refractory B-ALL, including those with high-risk features. Patients who received CAR T-cell therapy followed by consolidative allo-HSCT had **better leukemia-free survival (LFS)** and **overall survival (OS)** than those who received CAR T-cell therapy alone. |
| 20 | Pasquini et. Al. 2020 | Canada | 511 | 241 | 170 | CAR T-Cells | 511 | N/A | N/A | Single Arm | 13.4 | 13.2 | CAR T-Cells | 1 | CD19 | 4-1BB | CART | High | The real-world evidence of Tisagenlecleucel for pediatric ALL and adult NHL shows that its efficacy is comparable to pivotal trial results, with high rates of complete remission and manageable toxicity |
| 21 | Gardner et. Al. 2017 | USA | 45 | 23 | 22 | CAR T-Cells | 45 | N/A | N/A | Single Arm | 28 | 12.2 | CAR T-Cells | 1 | CD19 | 4-1BB | CART + HSCT | High | The main finding of this study was that **CD19 CAR T-cell therapy** demonstrated an **89% MRD-negative complete remission rate** in children and young adults with relapsed/refractory B-lineage acute lymphoblastic leukemia. The study found the therapy to be feasible with an acceptable toxicity profile, including cytokine release syndrome and neurotoxicity. |
| 22 | Leahy et. Al. 2022 | USA | 231 | 134 | 97 | CAR T-Cells | 231 | N/A | N/A | Single Arm | 24.7 | 12 | CAR T-Cells | 2 | CD19 | 4-1BB | CART | High | The main finding of the study is that **CD19-directed CAR T-cell therapy** is effective in achieving durable remissions in children and young adults with relapsed/refractory B-ALL, regardless of cytogenetic risk |
| 23 | Curran et. Al. 2019 | USA | 25 | 12 | 13 | CAR T-Cells | 25 | N/A | N/A | Single Arm | 28.6 | 13.5 | CAR T-Cells | 1 | CD19 | CD28 | CART + HSCT | High | The study found that **CD19-specific CAR T-cell therapy** for pediatric/young adult R/R B-ALL was **safe and feasible**, with a high response rate (75%) and manageable toxicity |
| 24 | Yan et. Al. 2024 | China | 36 | 18 | 18 | Blinatumomab bridging CAR-T | 18 | N/A | N/A | Single Arm | 8.13 | 43.5 | CD19 | 1 | CD19 | CD19 | CART + HSCT | High | sThe study demonstrated that blinatumomab bridging CAR-T therapy significantly improves the efficacy and safety in adult B-ALL patients, with better remission rates and lower cytokine release syndrome compared to CAR-T therapy alone. |
| 25 | Levine et. Al. 2021 | USA | 137 | 72 | 65 | CART | 137 | N/A | N/A | Single Arm | 24 | 12 | Tisagenlecleucel | 3.3 | CD19 | 4-1BB | CART + HSCT | High | The study found that **tisagenlecleucel** is effective for treating relapsed or refractory **B-cell acute lymphoblastic leukemia** (B-ALL) with common adverse effects including **CRS** and **neurologic events**, which were manageable with interventions like tocilizumab and corticosteroids |
| 26 | M. Myers et. Al. 2025 | USA | 19 | 13 | 4 | CART22-65s | 19 | N/A | N/A | Single Arm | 38 | 16.1 | CART22-65 | 1 | CD19 | 4-1BB | CART + HSCT | High | CART22-65s induced remissions in 74% of patients with highly refractory B-ALL, with a favorable safety profile and low rates of severe toxicity. However, remission durability was limited without consolidative HSCT |
| 27 | Zhao et. Al. 2021 | China | 105 | 58 | 42 | CART | 27 | Chemotherapy | 78 | Single Arm | 49 | 13 | CD19 | 2 | CD19 | 4-1BB | CART+ HSCT | High | The main finding indicated that **CART therapy followed by HSCT** for relapsed/refractory B-ALL resulted in comparable long-term outcomes (LFS and OS) to **chemotherapy followed by HSCT**, despite more advanced disease in the CART group |
| 28 | Hu et. Al. 2022 | China | 153320 | 77880 | 75440 | CART | 153320 | N/A | N/A | Observational Cohort | 348 | 9 | CART | 2 | CD19 | 4-1BB | CART + HSCT | High | The main finding is that the global burden of **acute lymphoblastic leukemia (ALL)** has increased from 66,810 cases in 1990 to 153,320 in 2019. The incidence rate has risen by 1.29% globally. **East Asia** had the highest increase in cases, and the prognosis for older patients is generally poorer. |
| 29 | Ma et. Al. 2019 | China | 10 | 5 | 5 | CART | 10 | N/A | N/A | Single Arm | 8.7 | 6.5 | CART | 0.5 | CD19 | 4-1BB | CART + HSCT | High | The study demonstrated that CD19-targeted CAR-T cell therapy was an effective treatment for pediatric patients with relapsed/refractory B-ALL. Although 80% of patients achieved complete remission (CR), 40% relapsed, mostly due to loss of CD19 expression |
| 30 | Shang et. Al. 2025 | China | 116 | 63 | 53 | CART | 116 | N/A | N/A | Single Arm | 47.9 | 6 | CART | 4.03 | CD19 | 4-1BB | CART + HSCT | High | Pre-infusion disease burden (such as MRD ≥ 1%) was associated with worse long-term survival outcomes (OS and EFS) and increased toxicity following anti-CD19 CAR T-cell therapy |
| 31 | Shah et. Al. 2021 | USA | 50 | 40 | 10 | CART | 50 | N/A | N/A | Single Arm | 57 | 13.5 | CD19 | 1 | CD19 | CD28 | CART+ HSCT | High | The study demonstrated that autologous CD19.28z-CAR T-cell therapy followed by consolidative alloHSCT (Hematopoietic Stem Cell Transplantation) can provide durable disease control and long-term survival in children and young adults with relapsed or refractory B-ALL |
| 32 | Maude et. Al. 2018 | USA | 75 | 42 | 33 | Tisagenlecleucel | 75 | N/A | N/A | Observational Cohort | 13.1 | 11 | CART | 3.1 | CD19 | 4-1BB | CART | High | The study demonstrated **high remission rates** and **durable remissions** with **Tisagenlecleucel** in pediatric and young adult patients with **relapsed or refractory B-cell ALL**. The **6-month overall survival** rate was **90%**, and **81% of patients** achieved complete remission |
| 33 | Roddie et. Al. 2024 | UK | 153 | 66 | 61 | Obecabtagene autoleucel | 127 | N/A | N/A | Single Arm | 21.5 | 47 | CART | 10 | CD19 | 4-1BB | CART | High | Obe-cel resulted in a high incidence of durable response among adults with relapsed or refractory B-cell ALL, with a low incidence of grade 3 or higher immune-related toxic effects. |
| 34 | Lee et. Al. 2015 | USA | 21 | 13 | 8 | CAR T-Cells | 21 | N/A | N/A | Single Arm | 10 | 15 | CD19 | 1 | CD19 | CD28 | CART + HSCT | High | The study found that **CD19-CAR T-cell therapy is feasible, safe**, and effective in treating **refractory B-cell acute lymphoblastic leukaemia (B-ALL)** in children and young adults, with **complete response rates of 66.7%** and **MRD-negative complete responses in 60% of patients** |
| 35 | Fry et. Al. 2018 | USA | 21 | 17 | 5 | CD22-CAR T | 21 | N/A | N/A | Single Arm | 6 | 15 | CD22-CAR | 1 | CD22 | 4-1BB | CART | High | The study demonstrated that CD22-CAR T-cell therapy is effective for patients with relapsed/refractory B-ALL, including those previously treated with CD19-directed immunotherapy. However, relapses occurred due to diminished CD22 expression . |
| 36 | Minnema et. Al. 2024 | Netherlands | 58 | 29 | 29 | Brexucabtagene autoleucel | 43 | N/A | N/A | Single Arm | 26.8 | 26 | CARt | 1 | CD22 | 4-1BB | CART + HSCT | High | The study concluded that brexu-cel showed clinical benefit with an improved overall survival (OS) compared to historical standard-of-care therapies |
| 37 | He et. Al. 2025 | China | 21 | 9 | 12 | CART | 21 | N/A | N/A | Single Arm | 20.4 | 29 | CART | 1 | CD19 | CD28 | CART | High | The study found that the third-generation 1928zT2 CAR-T cells provided high response rates and manageable safety in patients with CNS involvement, demonstrating durable remissions and good overall survival |
| 38 | Dai et. Al. 2020 | China | 6 | 4 | 2 | CD19 CART | 6 | N/A | N/A | Single Arm | 11 | 17 | CART | 1.7 | CD19 | 4-1BB | CART + HSCT | High | Bispecific CD19/CD22 CAR T-cell therapy showed feasibility, safety, and potent anti-leukemic activity in patients with relapsed/refractory B-ALL. Relapse occurred in some patients, mainly due to antigen escape |
| 39 | Pan et. Al. 2019 | China | 34 | 20 | 14 | CD22-CAR T | 34 | N/A | N/A | Single Arm | 14 | 10 | CD22-CAR | 7.5 | CD22 | 4-1BB | CART + HSCT | High | The study demonstrated that **CD22 CAR T-cell therapy** showed **high efficacy** in inducing remission in **r/r B-ALL** patients, who had failed prior CD19 CAR T-cell therapy. The therapy resulted in **80% complete remission (CR)** in evaluable patients. A subsequent allogeneic hematopoietic stem cell transplantation (HSCT) improved long-term survival, leading to a **1-year leukemia-free survival rate of 71.6%**. |
| 40 | Maude et. Al. 2021 | UK | 15 | 9 | 6 | CD19 CART | 15 | N/A | N/A | Single Arm | 28 | 8 | CD19 | 0.3 | CD19 | 4-1BB | CART | High | The study found that **AUTO3** targeting both **CD19** and **CD22** showed promising safety and efficacy with a **60% survival rate** at **12 months**. However, it highlighted that **long-term persistence of CAR T-cells** was a challenge . |
| 41 | Ceppi et. Al. 2018 | USA | 21 | 16 | 5 | CD19 CART | 21 | N/A | N/A | Single Arm | 12 | 13 | CD19 | 1.2 | CD19 | 4-1BB | CART | High | The study found that a small change in the CAR T-cell manufacturing process led to longer persistence of CAR T-cells and better remission rates. However, the risk of severe neurotoxicity (e.g., fatal cerebral edema) was noted as a potential concern with the effector-driven phenotype of the SCRI-CAR19v2. |
| 42 | Bufalo et. Al. 2019 | Italy | 17 | 9 | 8 | CD19 CART | 17 | N/A | N/A | Single Arm | 18 | 15 | CD19 | 3 | CD19 | 4-1BB | CART | High | The study suggests that iC9-CD19-CAR T-cell therapy is feasible, safe, and highly effective in treating highly resistant and relapsed BCP-ALL, with no major life-threatening toxicities and high rates of complete remission (CR). |
| 43 | Dourthe et. Al. 2019 | France | 55 | 28 | 27 | Tisagenlecleucel | 41 | N/A | N/A | Single Arm | 7.2 | 18.2 | CART | 2 | CD19 | 4-1BB | CART | High | The study confirmed the efficacy of CTL019 in heavily pretreated patients with refractory or relapsed B-ALL. High response rates were observed, with persistent remissions and potential for cure. Toxicity was manageable with appropriate monitoring and treatment. |

Table S2. New ottowa Scale for Risk of Bias

| Sr No | Author Name | **Representativeness of the Exposed** | **Selection of the Non-Exposed** | **Ascertainment of Exposure** | **Outcome Negative at Start** | **Comparability** | **Assessment of Outcome** | **Enough Follow-Up Period** | **Adequacy of Follow Up** | **Evaluation Criteria** | **Overall** |
| --- | --- | --- | --- | --- | --- | --- | --- | --- | --- | --- | --- |
| 12 | Hiramatsu et. Al. 2019 | 1 | 1 | 1 | 1 | 1 | 1 | 1 | 1 | 0 | 7 |
| 13 | Talleur et. Al. 2019 | 1 | 1 | 1 | 1 | 1 | 1 | 1 | 1 | 0 | 7 |
| 14 | Amrolia et. Al. 2019 | 2 | 1 | 1 | 1 | 1 | 1 | 1 | 1 | 0 | 8 |
| 15 | Shen et. Al. 2020 | 1 | 1 | 1 | 1 | 0 | 2 | 2 | 1 | 0 | 8 |
| 16 | Huynh et. Al. 2017 | 2 | 1 | 1 | 1 | 1 | 1 | 1 | 1 | 0 | 7 |
| 17 | Shah et. Al. 2025 | 1 | 1 | 1 | 1 | 0 | 1 | 1 | 1 | 0 | 7 |
| 18 | Wayne et. Al. 2023 | 1 | 1 | 1 | 1 | 1 | 1 | 1 | 1 | 1 | 8 |
| 19 | Zhang et. Al. 2020 | 1 | 1 | 0 | 0 | 0 | 1 | 1 | 1 | 1 | 6 |
| 20 | Pasquini et. Al. 2020 | 1 | 1 | 1 | 0 | 1 | 1 | 1 | 1 | 1 | 8 |
| 21 | Gardner et. Al. 2017 | 1 | 1 | 1 | 1 | 1 | 1 | 1 | 1 | 0 | 7 |
| 21 | Leahy et. Al. 2022 | 1 | 1 | 1 | 1 | 1 | 1 | 1 | 1 | 0 | 7 |
| 22 | Curran et. Al. 2019 | 1 | 1 | 1 | 1 | 1 | 1 | 1 | 1 | 1 | 8 |
| 23 | Yan et. Al. 2024 | 1 | 1 | 1 | 1 | 1 | 1 | 1 | 1 | 1 | 8 |
| 24 | Levine et. Al. 2021 | 1 | 1 | 1 | 1 | 1 | 1 | 1 | 1 | 1 | 7 |
| 25 | M. Myers et. Al. 2025 | 1 | 1 | 1 | 1 | 1 | 1 | 1 | 1 | 1 | 1 |
| 26 | Zhao et. Al. 2021 | 1 | 1 | 1 | 1 | 1 | 1 | 1 | 1 | 1 | 7 |
| 27 | Hu et. Al. 2022 | 1 |  | 1 | 1 | 1 |  | 1 | 1 | 1 | 1 |
| 28 | Ma et. Al. 2019 | 1 | 1 | 1 | 1 | 1 | 1 | 1 | 1 | 1 | 8 |
| 29 | Shang et. Al. 2025 | 1 | 1 | 1 | 1 | 1 | 1 | 1 | 1 | 1 | 7 |
| 30 | Shah et. Al. 2021 | 1 | 1 | 1 | 1 | 1 | 1 | 1 | 1 | 1 | 7 |
| 31 | Maude et. Al. 2018 | 1 | 0 | 1 | 1 | 0 | 1 | 1 | 1 | 0 | 6 |
| 32 | Roddie et. Al. 2024 | 1 | 1 | 1 | 1 | 0 | 1 | 1 | 1 | 1 | 7 |
| 33 | Lee et. Al. 2015 | 1 | 0 | 1 | 1 | 0 | 1 | 0 | 1 | 1 | 5 |
| 34 | Fry et. Al. 2018 | 1 | 1 | 1 | 1 | 1 | 1 | 1 | 1 | 1 | 8 |
| 35 | Minnema et. Al. 2024 | 1 | 1 | 1 | 1 |  | 1 | 1 | 1 | 1 | 8 |
| 36 | He et. Al. 2025 | 1 | 1 | 1 | 1 | 1 | 1 | 1 | 1 | 1 | 8 |
| 37 | Dai et. Al. 2020 | 1 | 1 | 1 | 1 | 1 | 1 | 1 | 1 | 1 | 8 |
| 38 | Pan et. Al. 2019 | 1 | 1 | 1 | 1 | 1 | 1 | 1 | 1 | 1 | 8 |
| 39 | Maude et. Al. 2021 | 1 | 1 |  | 1 | 1 | 1 |  | 1 | 1 | 8 |
| 40 | Ceppi et. Al. 2018 | 1 | 1 | 1 | 1 | 1 | 1 |  | 1 | 1 | 8 |
| 41 | Bufalo et. Al. 2019 | 1 | 1 | 1 | 1 | 1 | 1 |  | 1 | 1 | 8 |
| 42 | Dourthe et. Al. 2019 | 1 | 1 | 1 | 1 | 1 | 1 | 1 | 1 | 1 | 8 |
